# Risk of evolution driven population-wide emergence of mpox: the paradoxic effect of moderate interventions

**DOI:** 10.1101/2024.11.26.24317969

**Authors:** F. Nedényi, J. M. Benke, M. Szalai, G. Röst

## Abstract

The global mpox outbreak has recently been declared a public health emergency of international concern. In this paper, we investigate the spread of mpox primarily evolving and propagating within a core population before affecting the general population. A main public health concern is that through evolution, mpox gains the ability to widely spread in the entire population. We examine how effective various intervention strategies are in preventing this from happening. These non-pharmaceutical interventions include reducing disease transmission in the core population, in the general population, or in both. Our analysis encompasses the optimal timing for these interventions, considering the effects of early versus late intervention and the potential impact of different mutation patterns on disease spread. Our findings highlight that effective early intervention can be achieved with lower intensity, while delayed intervention requires stronger measures. Notably, our results reveal an intriguing phenomenon where moderate intervention could lead to worse outcome than no intervention. This counterintuitive outcome arises because moderate restrictions may prolong transmission chains within the core group, leading to more opportunities for the pathogen to acquire mutations resulting in higher transmission potential in the general population. Consequently, a comprehensive understanding of the role of the core group in disease dynamics and the mutation patterns is crucial for developing tailored and effective public health strategies.

## 1 Introduction

The mpox disease (formerly known as monkeypox) caused by the monkeypox virus MPXV, endemic to Africa for decades, was historically a localized concern, with minimal impact outside the continent. Consequently, research and development of vaccines and medical tools were sparse due to the lack of global urgency (see [27]). However, this changed dramatically after the outbreak in May 2022, making mpox modeling crucial for understanding and controlling the spread of the disease [23]. Before 2022, cases outside Africa were rare, and it was uncommon to identify confirmed or suspected mpox cases without direct travel links to endemic areas.

By May 26, 2022, mpox had spread to 23 non-endemic countries, predominantly affecting a close-knit subpopulation mainly consisting of men who have sex with men (MSM), totaling 257 cases without any reported deaths [23]. By June 27, 2022, the virus had expanded to 50 countries/territories across five WHO regions, with 3,413 laboratory-confirmed cases, primarily in non-endemic regions [25], and the total number of confirmed cases reached nearly one hundred thousand. In contrast, previous outbreaks were largely confined to Central and West African nations such as the Central African Republic, the Democratic Republic of the Congo (DRC), Nigeria, and the Republic of the Congo [7, 12].

In July 2022, the WHO declared mpox a public health emergency of international concern (PHEIC). Thanks to coordinated efforts among affected communities, public health authorities, and global partners, the outbreak was brought under control, leading to the termination of the PHEIC less than a year later. However, in September 2023, a new variant (clade 1b) emerged in the DRC and rapidly spread to four neighboring countries previously unaffected by mpox. This resurgence led to a new PHEIC declaration in August 2024. The virus has since spread beyond Africa, with cases reported in Sweden and Thailand. By the end of August 2024, the DRC alone reported over 18,000 suspected mpox cases and 615 deaths. Despite these challenges, the WHO maintains that mpox outbreaks can be controlled with appropriate measures [27].

Mpox is a viral zoonosis with symptoms similar to those of smallpox. It was first identified in 1958 in a laboratory monkey at the Statens Serum Institute in Copenhagen, Denmark, and the first human case was reported in a young child in the DRC in 1970 [23]. The virus, a double-stranded DNA virus from the Poxviridae family, is typically transmitted to humans through contact with biological fluids or lesions of an infected animal, often a rodent [10]. Human-to-human transmission occurs through contact with infected biological fluids, skin lesions, mucous membranes, or inhalation of contaminated particles [3]. The incubation period generally ranges from 6 to 13 days, but can extend from 5 to 21 days [23, 24].

The importance of this topic has inspired many researchers. Let us give a short overview on some of the related results. A deterministic model was created by Yuan et al. [28], where they modeled the spread of mpox in humans, considering potential animal hosts such as rodents (e.g., rats, mice, squirrels, chipmunks, etc.) and emphasizing their role in the transmission of the virus to high-risk groups such as gay and bisexual men. Peter et al. [20, 21] developed and analyzed a deterministic mathematical model for mpox. The outcomes of Khan et al. [14] reveal that the dynamic behavior of their proposed monkeypox 2022 system is mainly governed by some parameters that are precisely correlated with the noise intensities. Then, Endo et al. [9] calculated *R*_0_ for mpox transmission between men who have sex with men (a group that has seen a high number of cases in the 2022 outbreak), showing its value was substantially above 1 in this demographic group. For further publication related to the topic see Molla et al. [19].

The previous papers on mpox modeling do not examine the disease in the light of evolution towards widespread emergence. However, this approach has been successfully applied for other diseases in the past. In the following we give an overview of such research papers to motivate our approach on handling the mpox disease. Antia et al. [2] describe the dynamics and evolution of emerging diseases as a multi-type branching process. They calculated the extinction probabilities of the process numerically. A similar problem is discussed in the paper of Kollepara et al. [15], where they used a SIR-like model to argue that an initially dominant sexual transmission mode can be overtaken by casual transmission at later stages, even if the basic casual reproduction number is less than one. Their results highlight the risk of intervention designs developed solely based on the early dynamics of the disease. Alexander et al. [1] used a multi-type branching process to model the spread of an introduced pathogen evolving through several strains. Leventhal et al. [17] also worked out a continuous-time multi-type branching process, the central finding is that heterogeneity in contact networks, such as the presence of superspreaders, accelerates infectious disease spread in real epidemics. They show that the heterogeneity in contact structure, which facilitates the spread of a single disease, surprisingly renders a resident strain more resilient to invasion by new variants. Kucharski et al. [16] characterized the transmission potential of subcritical (*R <* 1) infections considering age stratification. Blumberg et al. [4, 5] also analyzed subcritical infections using branching processes to capture transmission heterogeneity, in the context of earlier monkeypox outbreaks. They evaluated surveillance requirements for detecting a change in the human-to-human transmission of mpox.

The 2023-2024 mpox outbreak presents distinct differences compared to the 2022-2023 outbreak, which predominantly spread in non-endemic countries through sexual contact among men who have sex with men. Two new variants have emerged, one of which now affects children; however, the transmission dynamics of these variants remain unknown. This current paper seeks to provide insight into effective interventions for managing these evolving outbreaks, contributing to global efforts to control and ultimately eradicate mpox. To achieve this, we develop a stochastic SEIR-type epidemiological model tailored to the 2022-2023 outbreak dynamics.

## 2 Methods

To model an mpox epidemic we take an approach that incorporates the properties of stochastic branching processes and also SEIR-type epidemiological models. To fit into the context of recent mpox outbreaks, we consider a population with the following structure: most of its individuals belong to the so-called *general population* (GP), while a small percentage belongs to a *core group* (CG). One could also consider multiple core groups, however, that is out of the scope of this article. As mentioned in the introduction, in recent years we have seen multiple different clades of the virus which have different transmission profiles. Moreover, due to many factors, such as environmental conditions, the outbreaks in the endemic African countries are very different from the ones we observe in non-endemic ones. Having a strong zoonotic transmission through rodents is a key difference, which, at the moment, is not considered in our model. Indeed, let us fix here that from here on, we are going to focus on the transmission profile that has been observed in non-endemic countries mainly during the 2022-2023 outbreak. (However, with slight modification of parameters one can easily fit the model to other situations.) Previous studies on mpox showed that the illness spreads mainly among men who have sex with men *(MSM)* [18, 22], *hence these individuals form the core group. The core group is close-knit in a sexual sense, so if one individual gets mpox, they are more likely to pass it within their group than in the general population*.

### 2.1 SEIR in a Given Group

*Consider a population of constant size N*. The SEIR approach means that at any time we keep track of the number of individuals in each disease state, which are:

- *S*, the number of susceptible individuals (those able to contract the disease);
- *E*, the number of exposed individuals (infected but not yet infectious);
- *I*, the number of infectious individuals (those capable of transmitting the disease);
- *R*, the number of recovered individuals (assumed to become immune).

Thus, *N* = *S*(*t*) + *E*(*t*) + *I*(*t*) + *R*(*t*) is unchanged over time.

Figure 1 shows the structure of the population. All of the individuals may contact each other, but the number of their contacts and the ways a potential transmission can happen differ. Susceptible individuals can be exposed by an infectious host, while recovered individuals can no longer spread or contract the illness. An exposed individual will become infectious after an exponential amount of time (with parameter *δ*).

**Figure 1:**
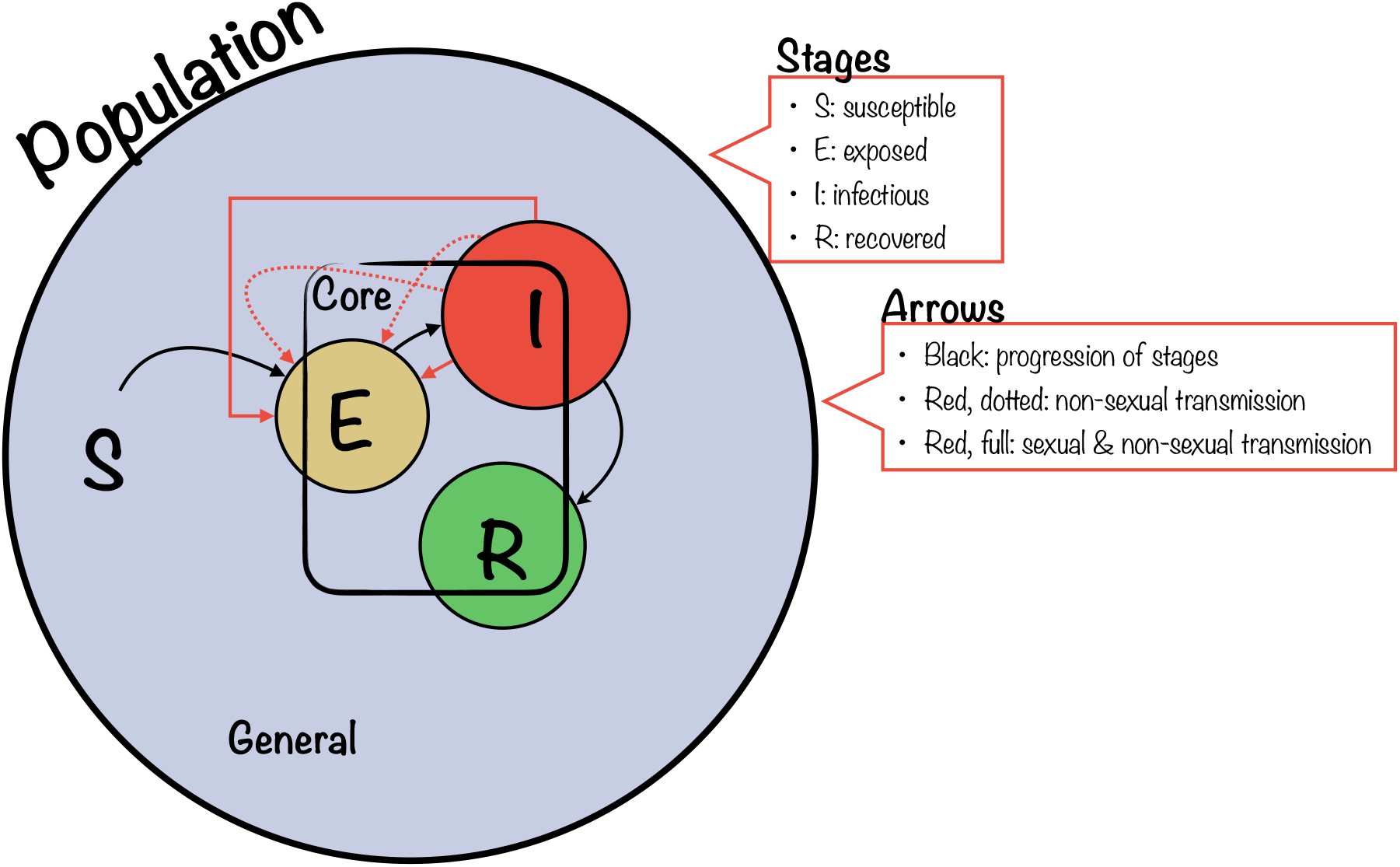
Dynamics among groups

An infectious individual recovers after an exponential amount of time (having parameter *γ*), but in the meantime it exposes susceptible individuals to the virus: again, exposure takes an exponential time with parameter *β*. This *β* parameter – which ultimately determines how many new sick individuals this current one creates – is specific to their type (general or core). The next section explains this parameter in depth.

### 2.2 Dynamics Among Groups

This subsection is about the structure and relationship among the general and the core group. Before explaining the specifics of our model, let us take a look at the the real-life background focusing on the non-endemic countries. As it has already been explained in the beginning of this section (Section 2), in non-endemic countries mpox tends to spread mainly within the high-risk MSM subpopulation. This is due to the fact that this particular illness mostly spreads through sexual contact and only secondarily via other types of contact including close contact, respiratory droplets, aerosols, and fomites. Moreover, the MSM community is known to have a closely interconnected social and sexual network, and this higher density of contact enables the virus to spread more readily. In our model we take into consideration two types of transmissions: sexual and non-sexual. We have the following assumptions that we established based on the history of the illness:

- The number of non-sexual contacts does not depend on the type of the individual.
- The non-sexual contacts of any individual have the same composition: the proportion of general individuals is the same as in the entire population. Thus if a transmission happens through nonsexual contact, then the probability that the newly exposed individual is a general one is the same as the proportion of general ones in the population.
- An individual only has sexual contact within its group.

These assumptions lead us to the following *β* values. Let us note here that there is no general *β*, this value, which is the number of expected offsprings of an infectious individual (per day), depends on the group of the infectious (lower index) and that of the potentially exposed (upper index) individual. We use the following notations:

- *p*^s^ and *p*^ns^ denote the probabilities of the sexual and non-sexual transmissions, respectively. This means that if a sexual or non-sexual contact occurs between two individuals, where one is able to infect the other, then this infection happens with probability *p*^s^ and *p*^ns^, respectively.
- *c*^ns^ denotes the number of expected non-sexual contacts per day.
- 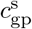and 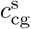denote the number of expected sexual contacts in the general population and in the core group, respectively, per day.
- *q*_gp_ and *q*_cg_ denote the proportion of the general and core group within the entire population, respectively.

Therefore, the 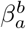 parameters, which, again, denote the expected number of people from group *b* infected by an infectious group *a* individual (per day) are the following:

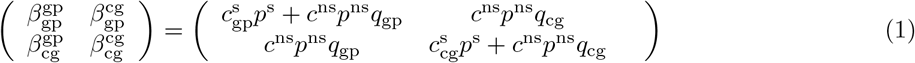

There is one additional aspect to be discussed, which is the mutation of parameter *p*^*ns*^. We know that the virus can evolve in the sense that it becomes more likely to spread via non-sexual ways. To mimic this property, each sick individual has a probability of non-sexual transmission. This value, *p*^*ns*^, is inherited from the individual who exposed the current one, but it may undergo mutation during this process. For more details on the actual mutation process see Section 2.3.

Here we also introduce the notations for the reproduction numbers of the individuals. This quantity, the expected number of people infected by an infectious individual during its infectious phase, is usually denoted by *R*_0_. However, for our study, we define this reproduction number in a specific way. We are particularly interested in the expected number of people infected by an infectious individual during its infectious phase within its own group. To this end, we introduce 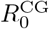 and 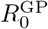, representing the reproduction numbers for individuals in the core group (CG) and the general population (GP) within their own group, respectively. These notations help us differentiate and track the reproductive rates within each group separately, which is crucial for understanding the dynamics of disease spread in structured populations.

### 2.3 Mutation Process

Each exposed individual inherits *p*^*ns*^, the probability of non-sexual transmission from the individual who exposed them to the virus, but this probability is subject to a mutation process in order to capture the evolution of the virus. In this paper we consider two types: the *d-step* and the *stochastic* mutation mechanism.

#### 2.3.1 *d*-Step Mutation Model

Based on the paper by Antia, et al. [2] one can introduce the following mutation process. When an individual becomes infected, the probability of non-sexual transmission is multiplied by a deterministic factor. This adjustment ensures that the mutation process reaches an 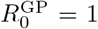 level after a specified number of steps, where 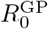 was introduced in Section 2.2. Our primary goal is to prevent the virus from spreading within the general population, which forms the majority of our total population. Therefore, with our previous notations from (1),

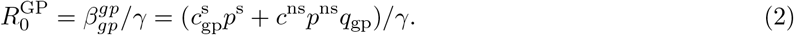

First of all one has to identify the initial 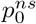 non-sexual transmission probability of the virus, and also the threshold 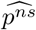, the probability of non-sexual transmission that results in 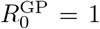 By Equation (2), this threshold probability is

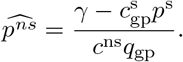

If we aim to define a mutation scheme that reaches 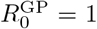 in *d* steps, then the multiplicative factor *r* to choose is

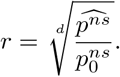

Then, to make sure that no transmission probability exceeds 1, we define the infected individual’s non-sexual transmission probability 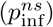 as

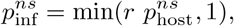

where 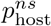 is the non-sexual transmission probability of the host.

To sum up, in this mutation process non-sexual transmission has an increasing pattern during a given number of mutation steps.

#### 2.3.2 Stochastic Mutation

Besides the previous *d*-step one, one can consider a different, but meaningful way to describe the mutation mechanism. In case of this method we assume that when an individual is infected by a host, the probability of non-sexual transmission is mostly copied from that of the host. There are two possible outcomes with given probabilities. In the first case, the infected individual inherits exactly the host’s probability of non-sexual transmission, meaning no mutation occurs. In the second case, the inherited probability is perturbed by defining a truncated normally distributed random variable centered around the host’s probability of non-sexual transmission. The truncation is symmetric, meaning the expected value of the perturbed probability equals the original value. The truncation interval is [max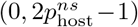, min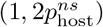]. The truncated normal distribution also has a parameter, the standard deviation, which determines the volatility of the mutation.

More precisely, let *p*^ir^ ∈ [0, 1] be the probability of inheritance. Then the distribution of 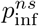 (depending on the respective probability of the host, 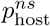) is given by:

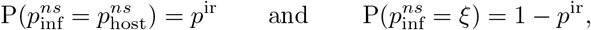

where the random variable *ξ* has a truncated normal distribution with mean 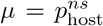, variance *σ*^2^, and truncation interval

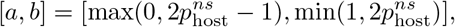

with the probability density function

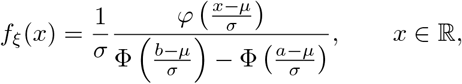

where *φ* is the probability density function of the standard normal distribution and Φ is its cumulative distribution function.

It is worth noting that the truncated normal distribution is the maximum entropy probability distribution for a fixed mean and variance when the corresponding random variable is constrained to a given finite interval, see [8]. Therefore, in this case, the mutation process is random, and the probability of non-sexual transmission can either increase or decrease in a mutation step, with a constant expectation.

### 2.4 The development of the population

In the previous sections, we constructed the mathematical model. Now, we describe the development of the population, specifically the simulation process. We use the Gillespie algorithm, which is a widely-used stochastic simulation method for modeling biochemical systems and population dynamics. The algorithm allows for the simulation of the system’s state changes over time by considering the probabilistic nature of events. However, due to the mutation mechanism in our model, the rates of these events vary dynamically [11].

Each time period leading to an action (such as becoming infectious, infecting a new individual, or recovering) follows an exponential distribution with a specific parameter. Consequently, the time before the next event in the population (whether an exposure, someone becoming infectious, or recovering) is determined by the minimum of several exponential random variables, which itself is exponentially distributed with a parameter equal to the sum of the individual rates. We then determine the type of action by further breaking down this aggregated parameter. Finally, we pinpoint the exact action by refining this breakdown even further. For a more detailed description of the process see Appendix C at the end of the paper. The entire Python code that produces all the outputs seen in this paper has been uploaded to GitHub. For details see Section D. We define two events that we consider as the termination of our simulation process. The first is *extinction*, which occurs when the epidemic ends, meaning there are no actively exposed or infectious individuals left in the entire population. Without these individuals, no new infection has a possibility, so the virus is extinct. The second is *evolution*, defined as the event when the average value of 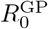 exceeds 1.

The values of the parameters that we used are summarized in Table 1. Parameters *p*^s^, *δ*, and *γ* were suggested by previous papers [6, 26]. The mutation related parameters *σ* and *d* have been set such that the baseline proportion of the evolution is around 50%. The number of initial infectious individuals *i* is equal to 10, because in this case the approximated probability that the process dies out immediately in the core group (approximately 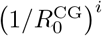) is negligible. The rest of the parameters were chosen to produce scientifically meaningful and accurate results.

**Table 1:**
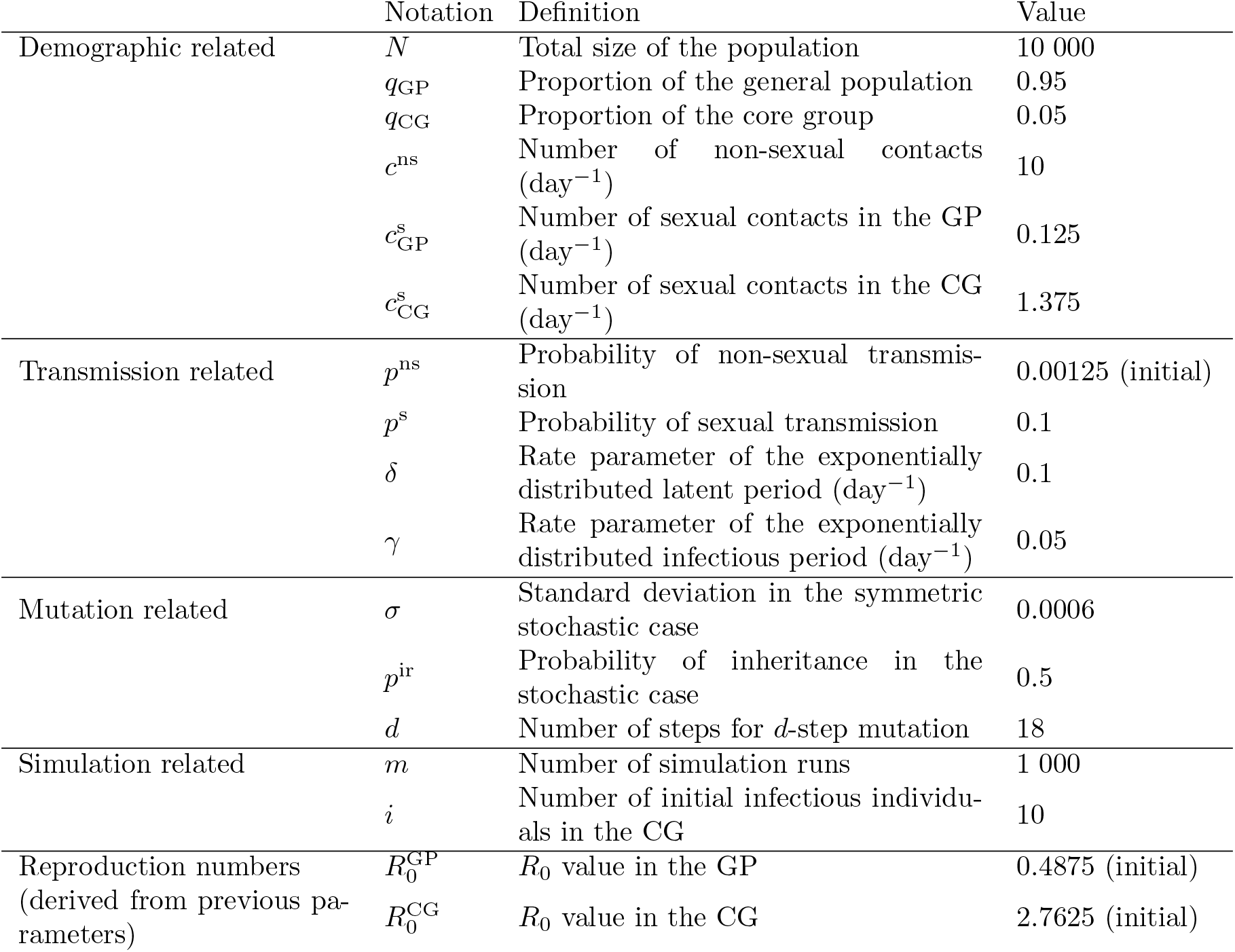
Parameter values.

|  | Notation | Definition | Value |
| --- | --- | --- | --- |
| Demographic related | $N$ | Total size of the population | 10 000 |
| | $q_{\text{GP}}$ | Proportion of the general population | 0.95 |
| | $q_{\text{CG}}$ | Proportion of the core group | 0.05 |
| | $c^{\text{ns}}$ | Number of non-sexual contacts (day<sup>-1</sup>) | 10 |
| | $c_{\text{GP}}^{\text{s}}$ | Number of sexual contacts in the GP (day<sup>-1</sup>) | 0.125 |
| | $c_{\text{CG}}^{\text{s}}$ | Number of sexual contacts in the CG (day<sup>-1</sup>) | 1.375 |
| Transmission related | $p^{\text{ns}}$ | Probability of non-sexual transmission | 0.00125 (initial) |
| | $p^{\text{s}}$ | Probability of sexual transmission | 0.1 |
| | $\delta$ | Rate parameter of the exponentially distributed latent period (day<sup>-1</sup>) | 0.1 |
| | $\gamma$ | Rate parameter of the exponentially distributed infectious period (day<sup>-1</sup>) | 0.05 |
| Mutation related | $\sigma$ | Standard deviation in the symmetric stochastic case | 0.0006 |
| | $p^{\text{ir}}$ | Probability of inheritance in the stochastic case | 0.5 |
| | $d$ | Number of steps for $d$-step mutation | 18 |
| Simulation related | $m$ | Number of simulation runs | 1 000 |
| | $i$ | Number of initial infectious individuals in the CG | 10 |
| Reproduction numbers<br>(derived from previous parameters) | $R_0^{\text{GP}}$ | $R_0$ value in the GP | 0.4875 (initial) |
| | $R_0^{\text{CG}}$ | $R_0$ value in the CG | 2.7625 (initial) |

## 3 Results

In the results section, we first examine the base scenario, where the virus is allowed to spread without any restrictions. Figures 2 and 3 show typical baseline scenarios for stochastic and *d*-step mutation with evolution on a single simulation. It is worth mentioning that for stochastic mutation the average 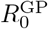 values have an increasing pattern, although the mutation is symmetric, meaning that the 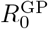 values can both increase and decrease. This phenomena can be explained by the evolutionary effect of the model.

**Figure 2:**
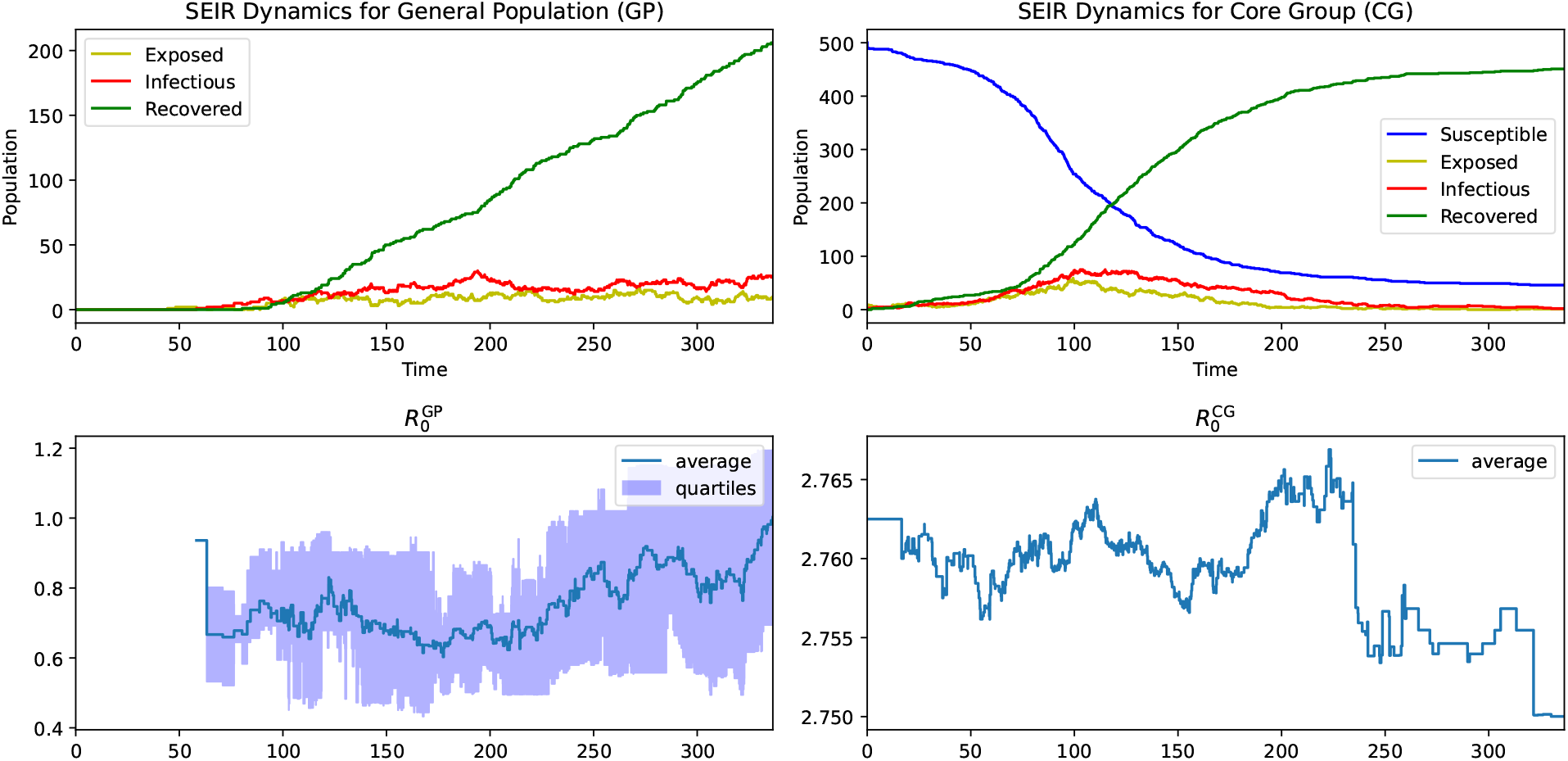
Baseline SEIR dynamics with 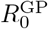and 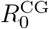 for **stochastic mutation**.

**Figure 3:**
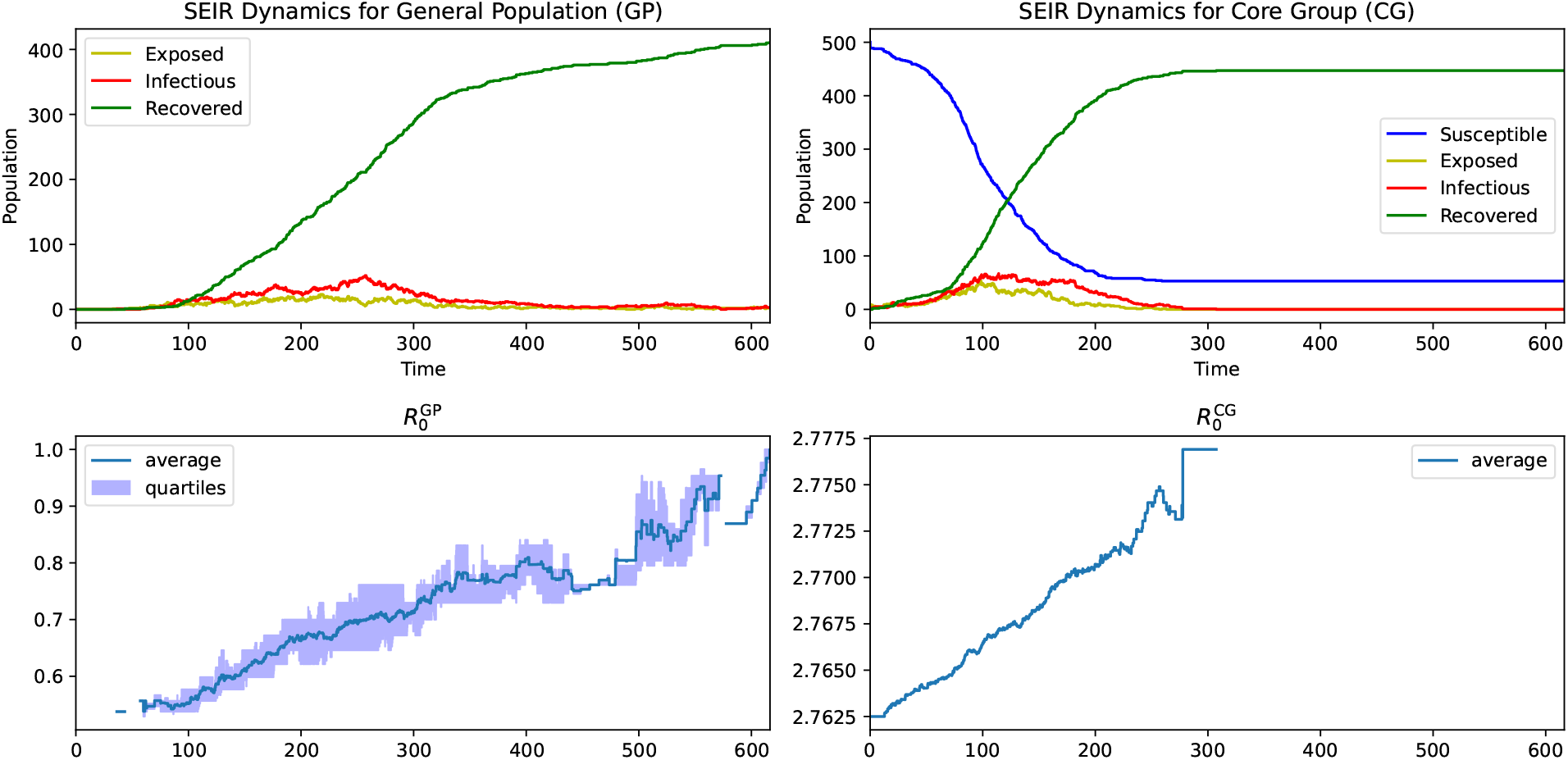
Baseline SEIR dynamics with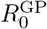 and 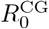 for *d***-step mutation**.

Following this, we explore various intervention strategies to understand their effectiveness. Non-pharmaceutical interventions are categorized in the following way.

1. Type of restrictions:
  - non-sexual restriction in the whole population;
  - sexual restriction in the core group.
2. Intervention is triggered when the proportion of infectious individuals is reaching the *intervention threshold* in:
  - the core group;
  - the general group; or
  - the entire population.

A specific restriction can be implemented by decreasing the number of the corresponding contacts (non-sexual or sexual) with a certain proportion (called *restriction level*), or decreasing the chance of transmission during a contact by personal protection measures, both leading to a reduction in the transmission rate. Let us note here that during our simulation study, when a sexual contact restriction is implemented, we only target the sexual contact number of the core group, not that of the general group. Finally, we analyze the proportion of trajectories where the virus evolves as previously described, focusing on cases where the average 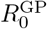 exceeds 1.

### 3.1 Analysis of Restriction Levels in Different Scenarios

We explored various scenarios to understand the impact of interventions on controlling the virus spread. Specifically, we focused on stochastic mutation with a restriction level of 50% (Figure 4), and *d*-step mutation with restriction levels of 50% (Figure 5) and 70% (Figure 6).

**Figure 4:**
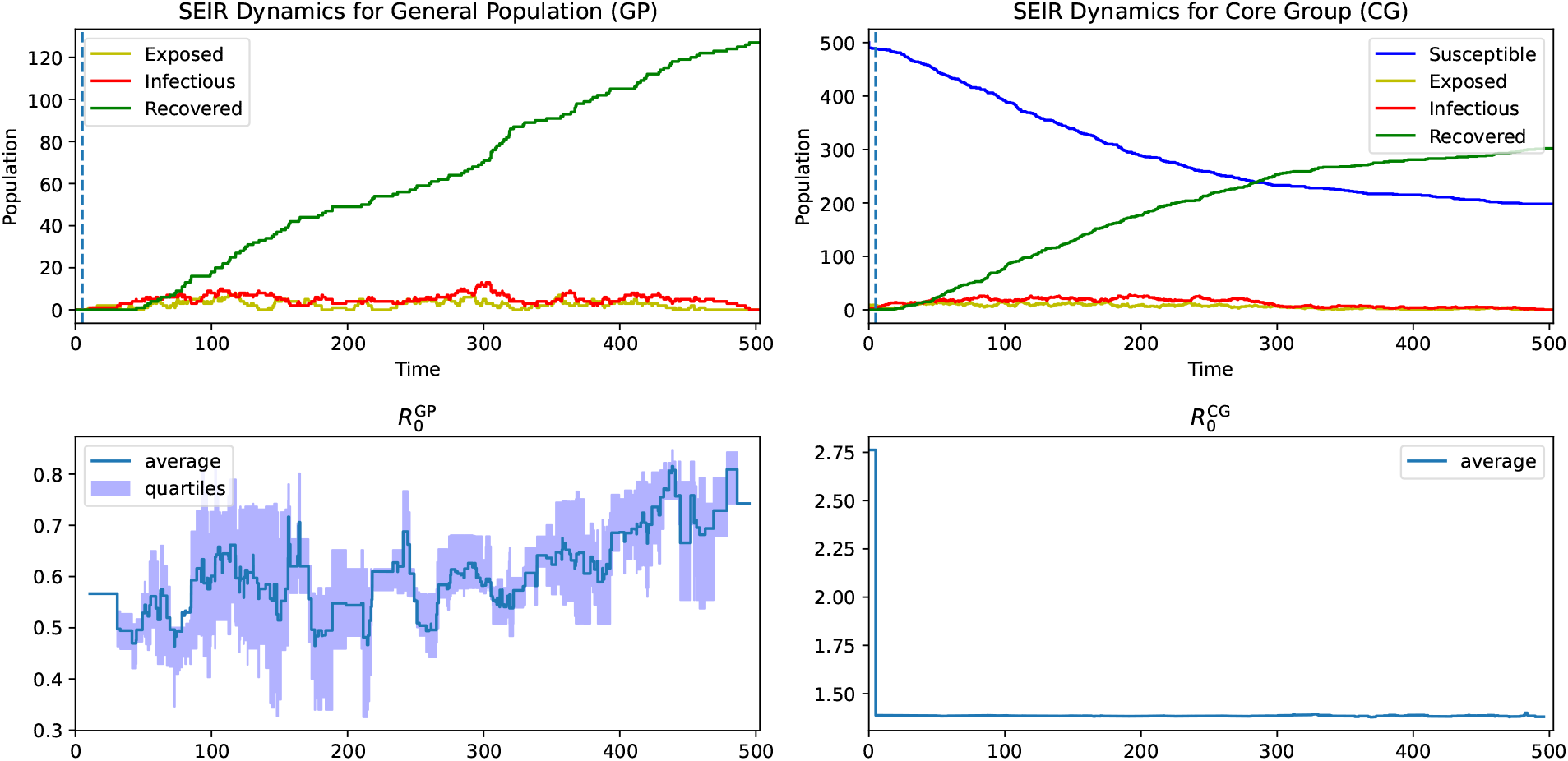
SEIR dynamics with **stochastic mutation**; restriction level 50%; sexual restriction in the core group; triggered by the core group; with intervention threshold 0.005.

**Figure 5:**
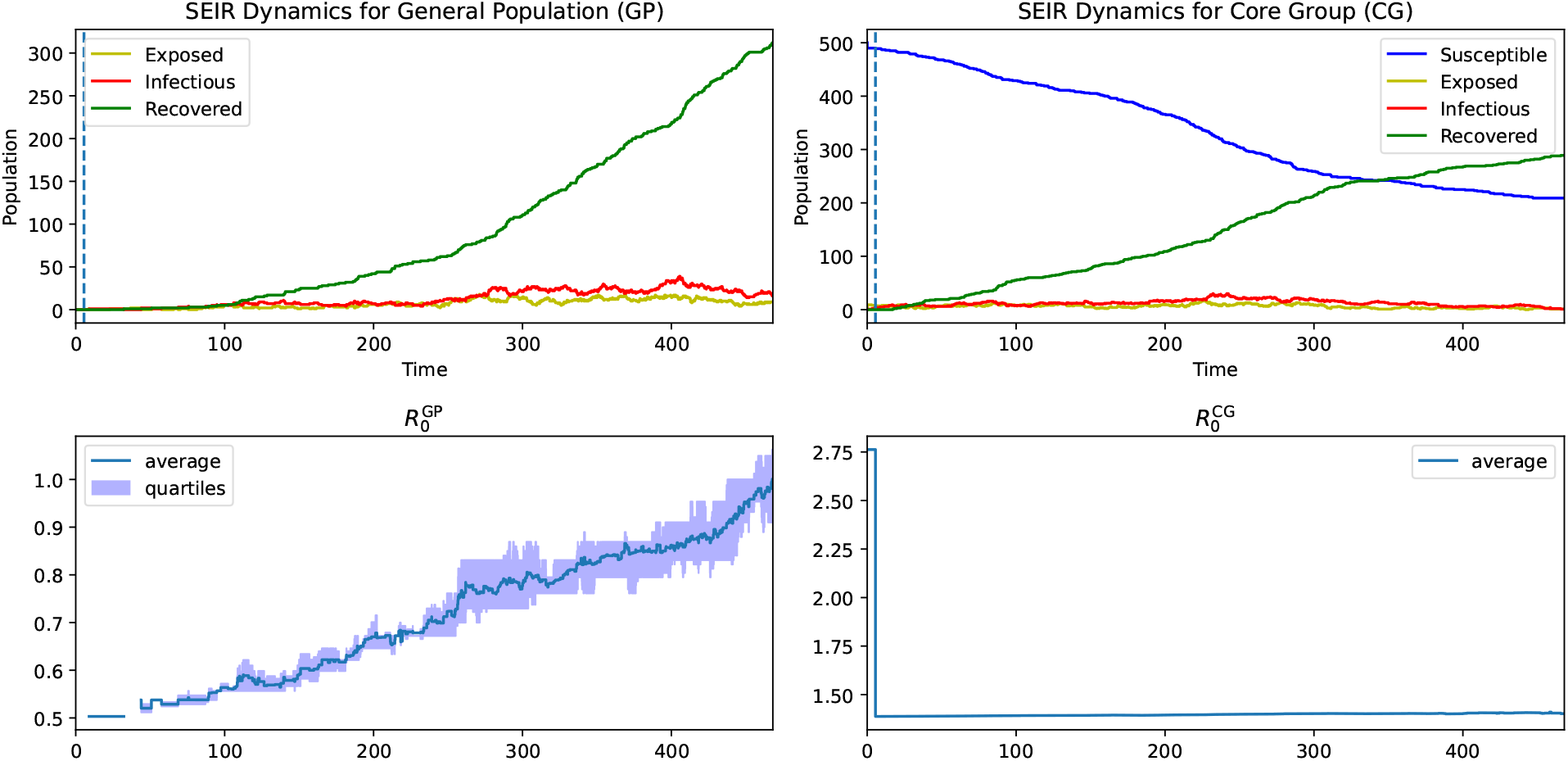
SEIR dynamics with *d***-step mutation**; restriction level 50%; sexual restriction in the core group; triggered by the core group; with intervention threshold 0.005.

**Figure 6:**
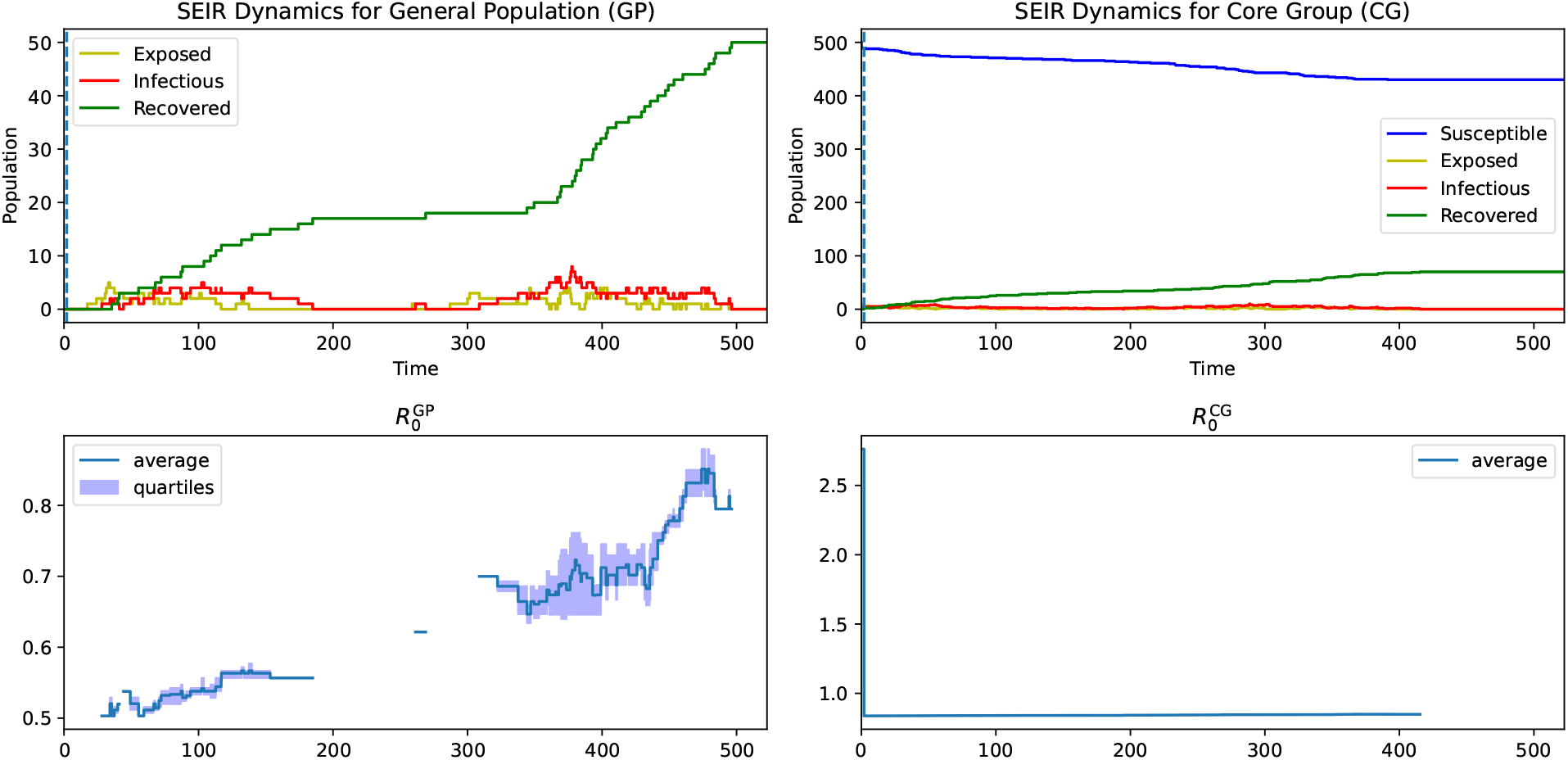
SEIR dynamics with *d***-step mutation**; restriction level 70%; sexual restriction in the core group; triggered by the core group; with intervention threshold 0.005.

In the stochastic mutation scenario (Figure 4), we applied a restriction level of 50% to the core group, reducing their sexual contact numbers by half. This resulted in the average 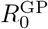 remaining below 1, indicating effective control of the virus spread.

For the *d*-step mutation scenarios (Figures 5 and 6), we applied restriction levels of 50% and 70%. To keep 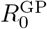 below 1, the model applying *d*-step mutation required more stringent restrictions (70%) compared to the stochastic model (50%). This suggests that *d*-step mutation, while more predictable, might necessitate stricter control measures to achieve the same level of epidemic control. Let us note here that this is expected as non-sexual transmission rate in case of stochastic mutation has a fixed expectation, however, the expectation strictly increases with each step for *d*-step mutation.

In summary, both stochastic and *d*-step mutation simulation results show that restricting sexual contacts within the core group effectively controls the spread of the virus. However, the level of necessary restriction differs. Indeed, the simulation applying stochastic mutation required a moderate restriction (50%), while the one with *d*-step mutation needed a more severe restriction (70%) to achieve similar results. This highlights the importance of tailored interventions based on the mutation type and spread dynamics to develop effective public health strategies for epidemic control.

### 3.2 Sensitivity Analysis Using Heatmap

The following heatmaps show the outcomes of different intervention strategies under both *d*-step and stochastic mutations. We focus on the effectiveness of these interventions in controlling the spread of the virus, particularly in maintaining the average 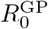 below 1.

These heatmaps illustrate the effectiveness of various intervention strategies under both *d*-step and stochastic mutations. In general, stricter restrictions (higher values of restriction level) result in a lower proportion of trajectories where the virus evolves to an 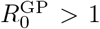. However, it is crucial to closely examine the specific circumstances under which this principle holds true, as well as the scenarios where this seemingly intuitive assumption may fail. This nuanced understanding helps in identifying the most effective intervention strategies tailored to the dynamics of the virus and the characteristics of the population.

Through sensitivity analysis, we aimed to identify the most effective combination of intervention type and timing to control the spread of the virus within the population. By examining various scenarios, we can better understand the impact of different restrictions and develop tailored strategies for epidemic control. The exact numbers of the simulation results can be found in the Appendix A. The heatmaps have been generated by a smoothing algorithm with bicubic interpolation. This technique is used to smooth the visual representation of the data, creating a more visually appealing gradient between data points. The bicubic interpolation works by considering the closest 4×4 neighborhood of known data points for each interpolated point. It then fits a cubic polynomial to these points, ensuring that the resulting image has continuous first and second derivatives, providing a smoother transition between colors in the heatmap.

### 3.3 Analysis of Transmission Chains

In this subsection we provide heatmaps and graphs to illustrate the effect of some of our restrictions on the length of the chain of transmission. On a chain of transmission we mean the following: the source is an individual who is infected when the simulation starts, its child nodes are the individuals infected by it, then their child nodes are the ones infected by them, and so on. The average length of transmission chain is calculated by taking the mean of the lengths of all paths from an initial infected individual (the source) to each leaf node in its transmission tree. This gives a measure of the typical chain length, accounting for both shorter and longer branches. As we will explain in Section 4, chain lengths are important in understanding how certain intervention strategies behave. In Figure 9 one can see the mean length of transmission chain for both *d*-step and stochastic mutation applying sexual restriction. The pattern is similar as in the case of the proportion of evolution, see the corresponding heatmaps in Figure 8. To illustrate the close connection between these quantities, we also present Figure 10. For a fixed intervention level (0.005) this plot shows the relationship between the average length of transmission chain and the proportion of evolution for both *d*-step and stochastic mutations. Let us note here that the plot for *d*-step mutation shows a very close connection between the two quantities, while for stochastic mutation, the two graphs are not as synchronized. The reason behind this is that for *d*-step mutation the place of an individual within the transmission chain determines its reproduction number, while for sexual mutation there is no direct connection as mutations can increase or decrease the probability of non-sexual transmission. Moreover, we plot transmission graphs in Figures 13, 14, 11 and 12. Each node in the graph represents an individual, the circles are in the core group, the boxes in the general population. The edges represent the infections. The number in each node is the length of the path from the source, so the number in a leaf node is the length of the corresponding transmission chain. A node in the general population is colored orange if the corresponding 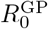 value is greater than 1. Meanwhile, a node in the core group is colored red if an individual in the general group having the same non-sexual transmission probability as the current core group individual would have an 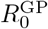value bigger than 1. In other words, if the core group individual exposed a general one who inherited its non-sexual transmission probability without any mutation, then the exposed individual would be a threat for causing an outbreak in the general population. Here we present four forests: a baseline and a restriction scenario for *d*-step and stochastic mutation each. Let us point out that in case of *d*-step mutations if at some point a node is colored orange or red, then all of its descendants are colored as well. Indeed, this type of mutation only increases the corresponding reproduction number. However, for the stochastic mutation an orange or red node can have non-colored descendants as this mutation can potentially decrease reproduction number as well.

## 4 Discussion

The key findings of our study carry implications for controlling the spread of diseases that evolve and spread within a host population with a core group structure.

From the heatmaps of the sensitivity analysis, we observe that non-sexual restrictions (Figure 7) are quite straightforward: the earlier and more severely we intervene, the better the outcomes in terms of controlling the virus spread. Note that due to the small differences in the proportions the effect of earlier or later intervention is not too visible on the heatmaps, but one can check the exact numbers in the Appendix A. However, for sexual restrictions in the core group, an interesting phenomenon is observed. While strong interventions are effective, moderate interventions can actually worsen the situation. (See red zone in Figure 8) This counterintuitive result occurs because it can sometimes be more advantageous to allow the virus to burn through the core group quickly rather than having a slower spread resulting in long transmission chains. Longer transmission chains lead to more mutations and higher transmission probabilities. If the virus is transmitted from the core group to the general group at a later phase of the transmission chain, it results in a higher proportion of evolved viruses with increased transmission capability. This explains why the corresponding heatmap regions have a middle section with higher numbers, indicating a worse outcome for moderate interventions compared to no intervention. One can read about this effect (longer chains resulting in a worse outcome) in [13], which describes that it may be necessary to intervene even if a virus is spreading slowly (*R*_0_ smaller than 1 or close to it), since allowing it to adapt can cause huge problems later on. Let us draw special attention to the heatmap corresponding to sexual restrictions based on the whole population in case of the *d*-step mutation model. The aggregate of many different effects shows on this plot (Top right heatmap of Figure 8). Here an intervention is initiated when a certain proportion of the entire population is infected, which is probably the closest to the real-life situation as it is not standard to report or know the sexual preference of an infected person. If the intervention threshold is low, meaning that a restriction is triggered in the early stages of the viral outbreak, most of the infected individuals come from the core group still. Then interventions behave similarly to when monitoring is done in CG directly, a weak (or no) restriction may be better than a moderate one. As the intervention threshold increases, this effect becomes less prominent and eventually disappears.

**Figure 7:**
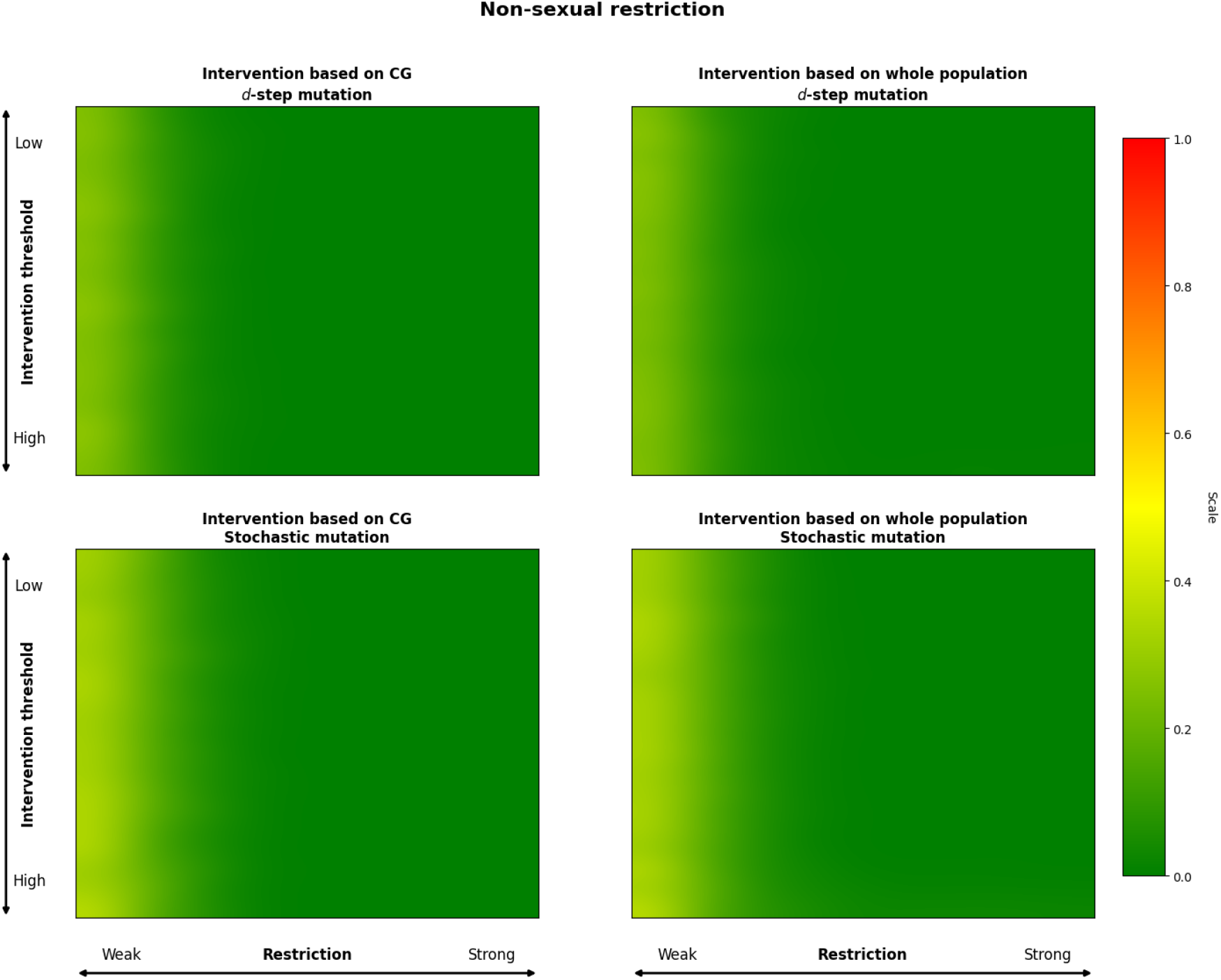
Heatmaps showing the proportion of trajectories where 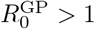 (which we call an evolution) for different intervention strategies relying on non-sexual restrictions. The top row represents *d*-step mutations, and the bottom row represents stochastic mutations. The left column interventions are based on the core group, and the right column interventions are based on the whole population.

**Figure 8:**
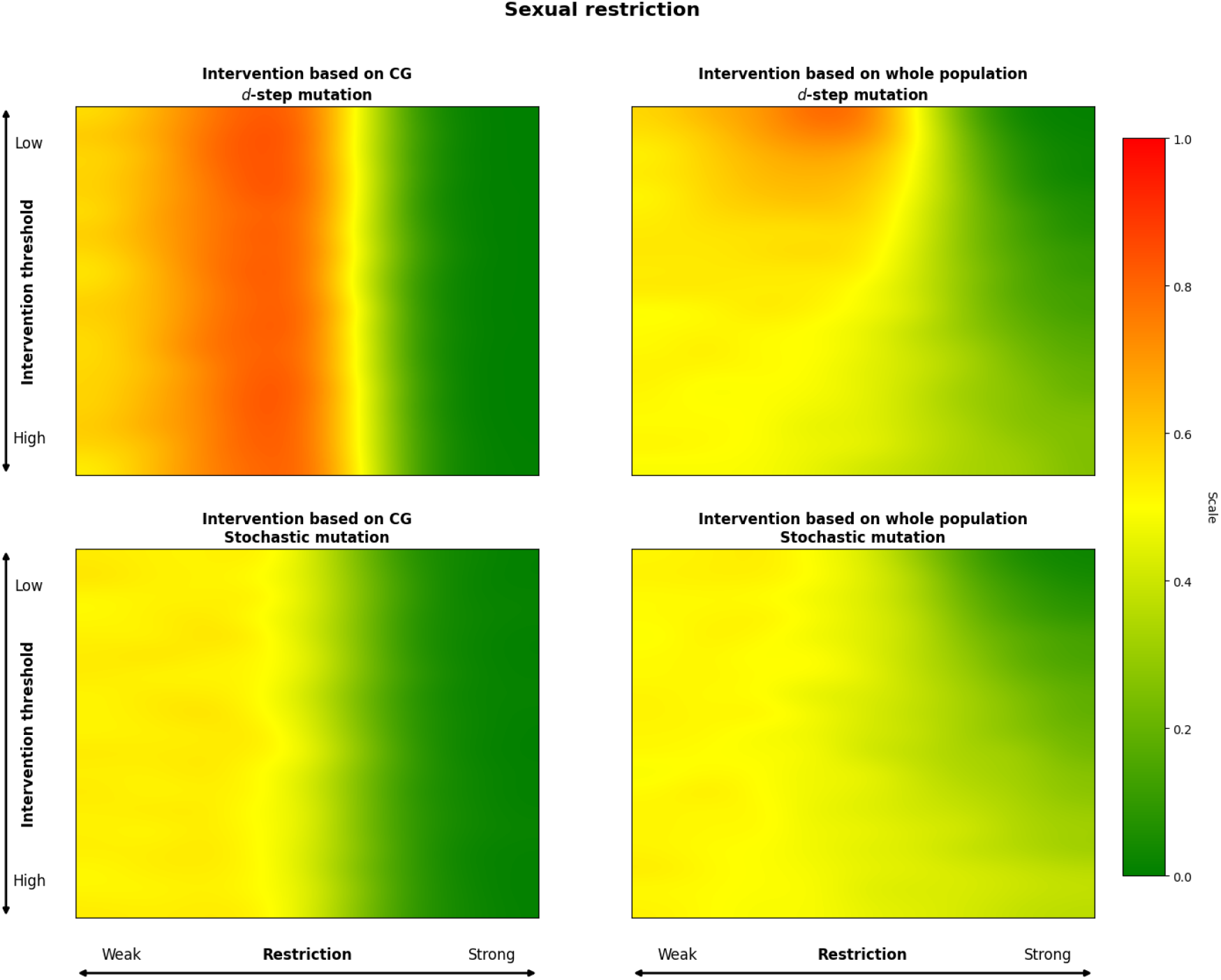
Heatmaps showing the proportion of trajectories where 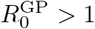 (which we call an evolution) for different intervention strategies relying on sexual restrictions. The top row represents *d*-step mutations, and the bottom row represents stochastic mutations. The left column interventions are based on the core group, and the right column interventions are based on the whole population.

One of the critical aspects we examined is the dependency between in-host selection and the transmissibility of the virus. Our results indicate that the strength of this dependency significantly influences the required intervention strategies. In scenarios with weak in-host selection, mitigation strategies, such as those observed in the stochastic mutation model, are sufficient. However, in cases of strong in-host selection, more aggressive suppression strategies are necessary, as seen in the case of the *d*-step mutation model.

The length of the transmission chain also plays a crucial role in the dynamics of virus spread and evolution. Longer transmission chains lead to more opportunities for the virus to mutate and acquire higher transmissibility through non-sexual contacts. This is particularly evident in the *d*-step mutation model, where longer chains result in eventually higher transmission probabilities due to accumulated mutations. See Figures 9 and 10, along with the transmission graphs in Figures 11 and 12 in Subsection 3.3 showing the strong relationship between evolution and chain length. This phenomenon underscores the importance of early intervention to shorten transmission chains and reduce the risk of highly transmissible virus strains emerging. For further reading on the impact of transmission chain length, see [13].

**Figure 9:**
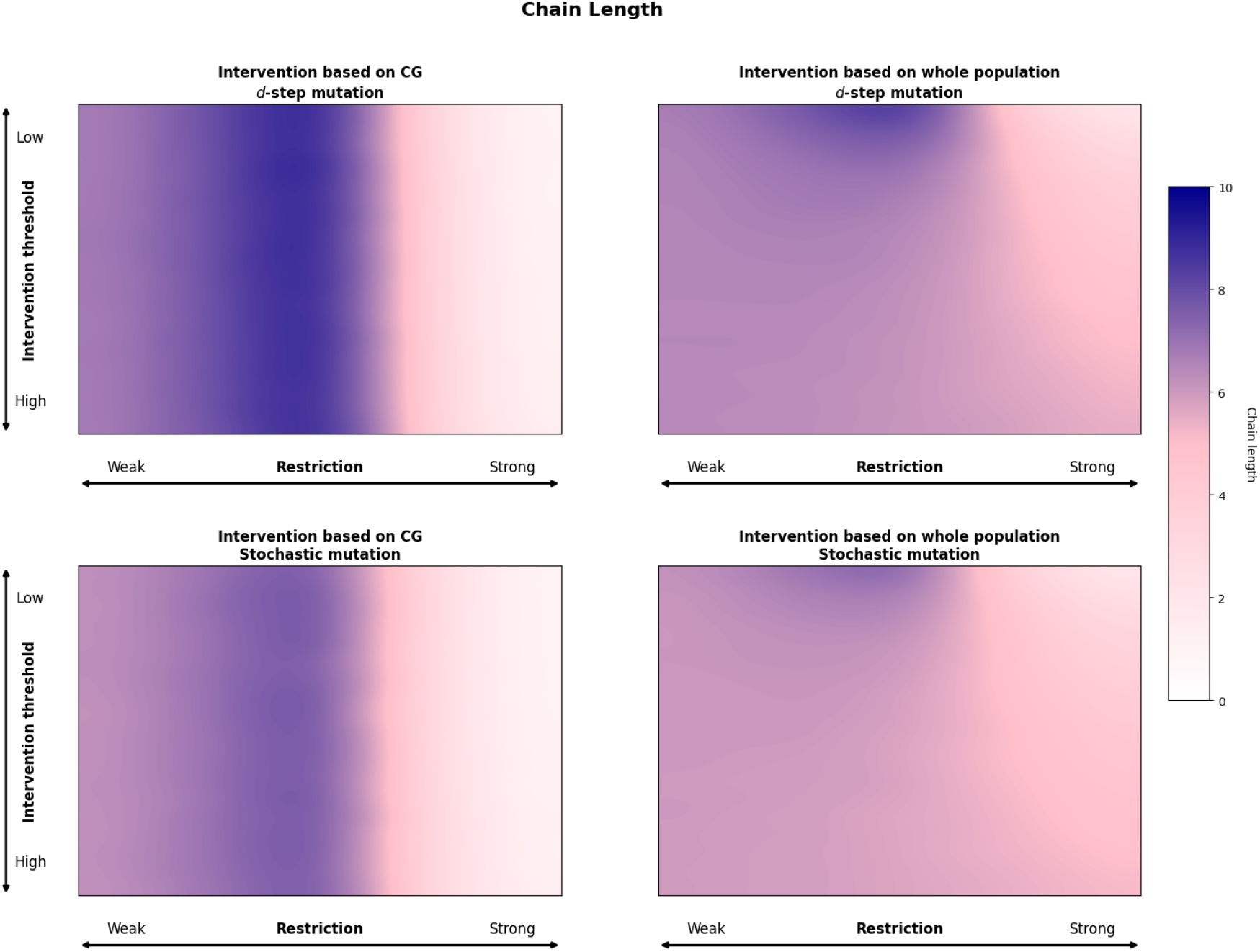
Heatmaps showing the average length of transmission chains. The left column interventions are based on the core group, and the right column interventions are based on the whole population.

**Figure 10:**
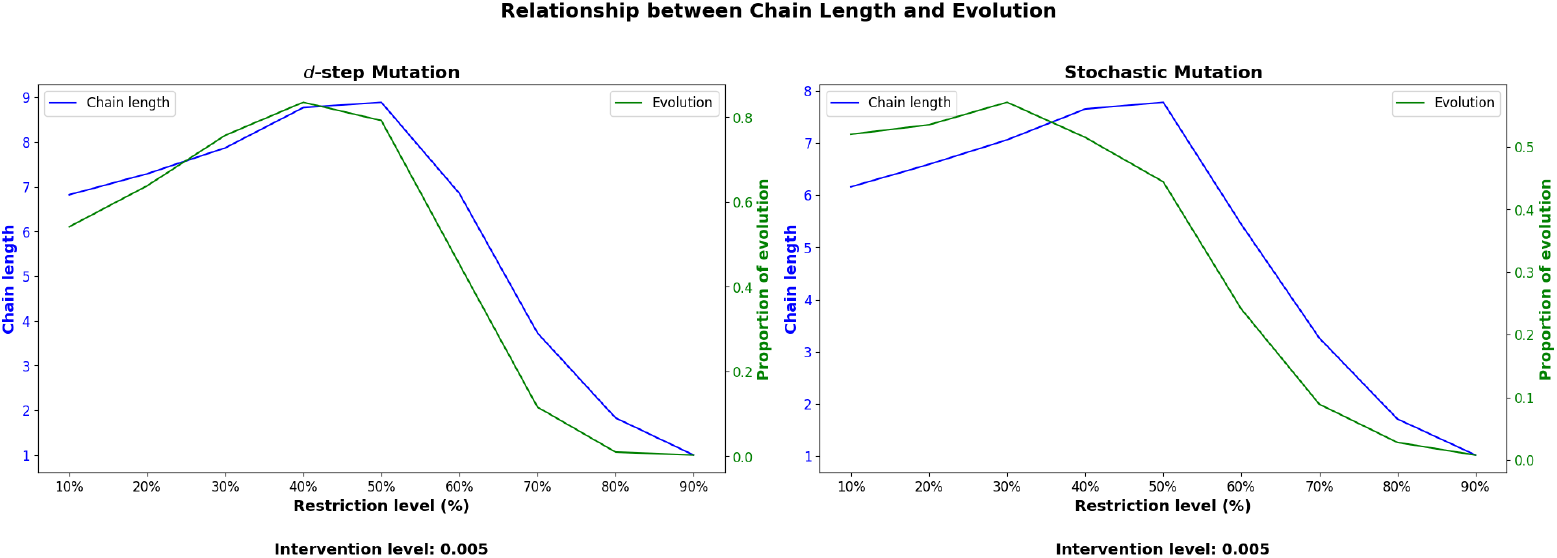
Graphs showing how for a fixed intervention level (0.005) the size of restriction effects the average length of transmission chain (blue function) and the proportion of evolution (green function). Note that scaling is different for the two variables.

**Figure 11:**
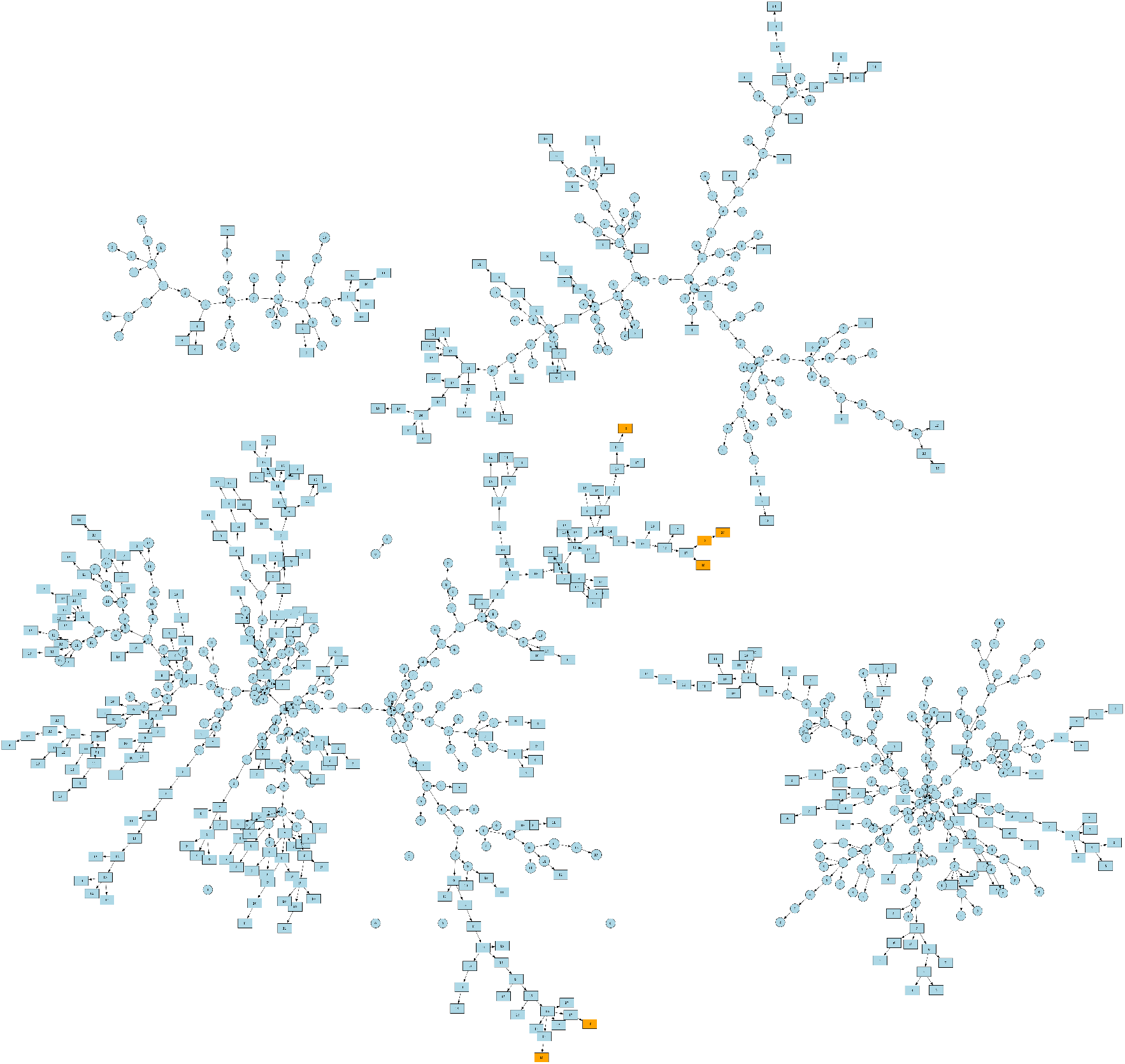
Baseline transmission graph for *d***-step mutation**. Mean of the transmission chain lengths is 7.87.

**Figure 12:**
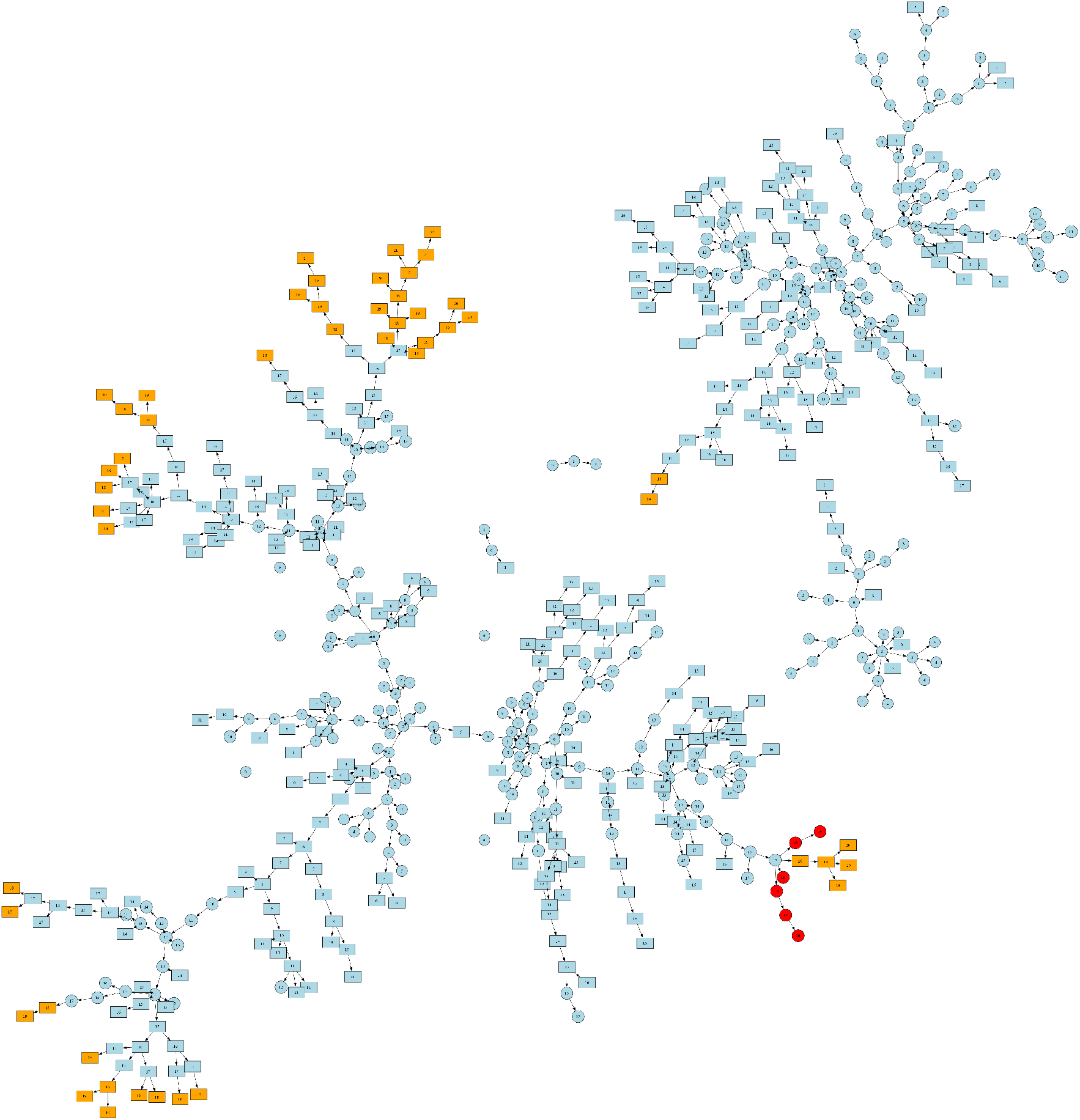
Transmission graph for *d***-step mutation**; restriction level 50%; sexual restriction in the core group; triggered by the core group; with intervention threshold 0.005. Mean of the transmission chain lengths is 11.16.

**Figure 13:**
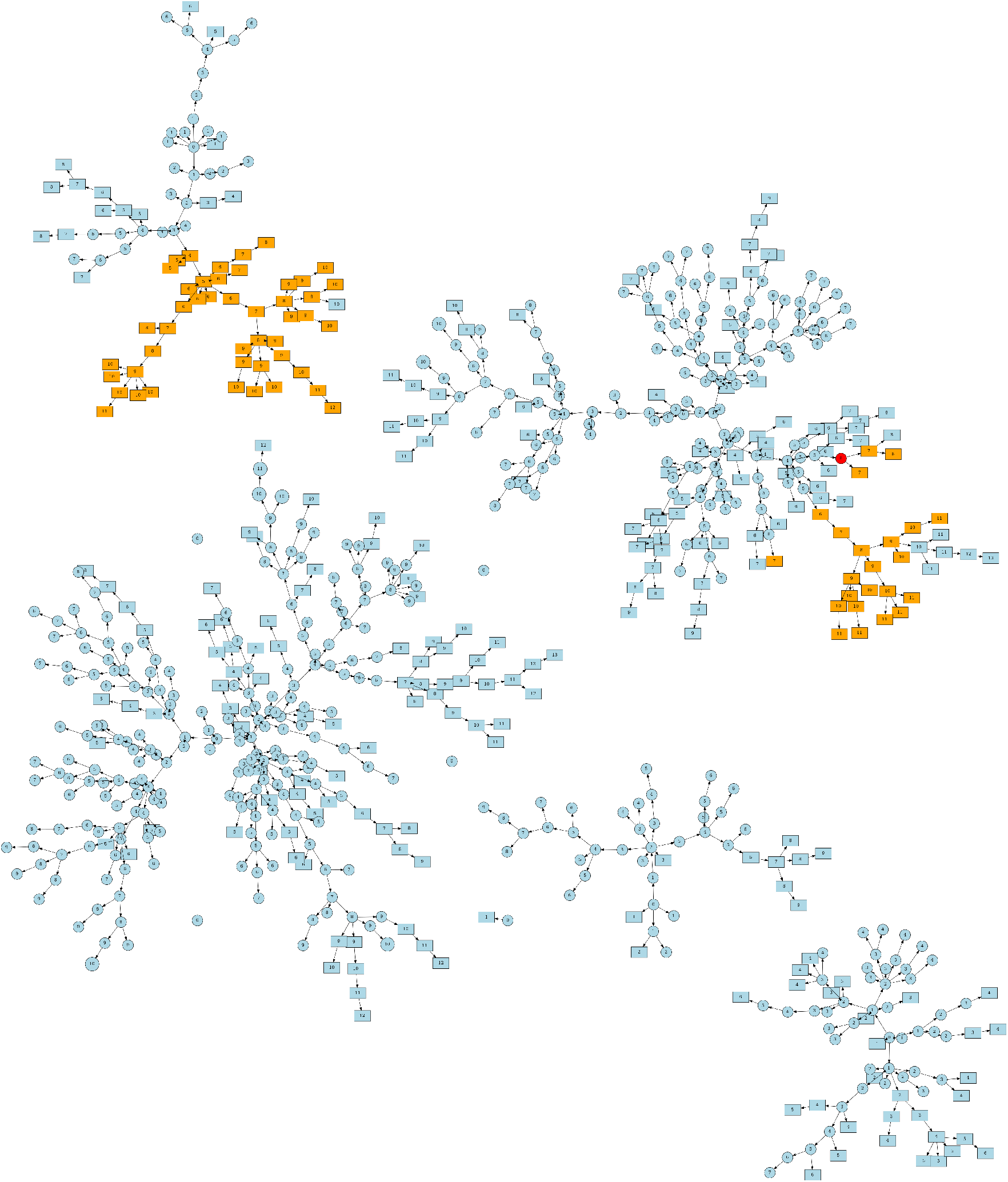
Baseline transmission graph for **stochastic mutation**. Mean of the transmission chain lengths is 6.03.

**Figure 14:**
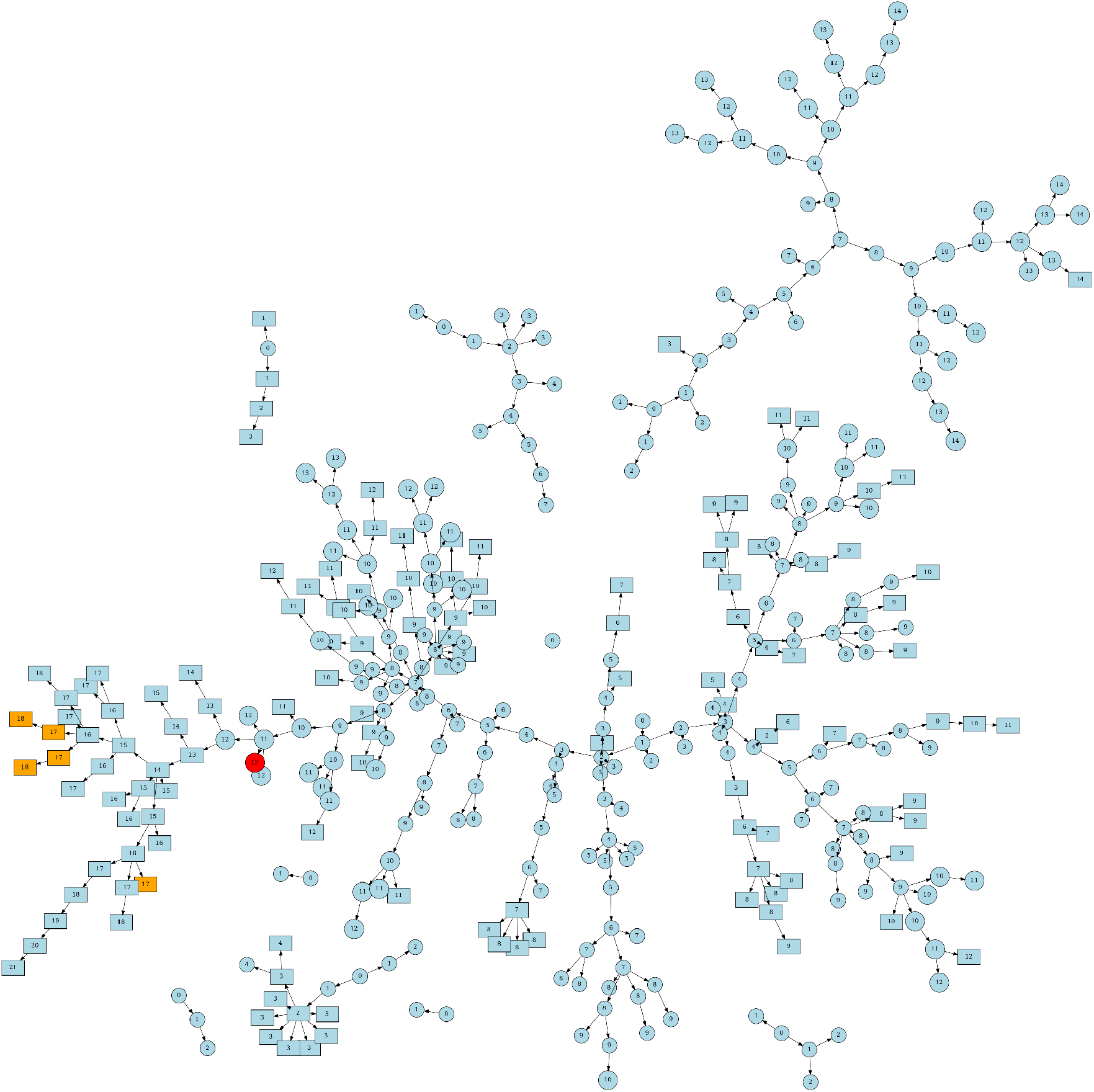
Transmission graph for **stochastic mutation**; restriction level 50%; sexual restriction in the core group; triggered by the core group; with intervention threshold 0.005. Mean of the transmission chain lengths is 8.74.

Future research can extend such investigations to other diseases with similarly structured populations, and expand the analysis by exploring multiple core groups and their interactions. Understanding the dynamics in these more complex scenarios will help refine our intervention strategies and improve our ability to control or mitigate the spread of infectious diseases.

## Data Availability

All data produced in the present work are contained in the manuscript.

https://github.com/HCEMMScientificComputingACF/Mpox

## Acknowledgement

The authors were supported by the National Research, Development and Innovation Office (NKFIH) in Hungary, grants RRF-2.3.1-21-2022-00006, TKP-2021-EGA-05, KKP 129877, 2022-2.1.1-NL-2022-00005. The project has received funding from the EU’s Horizon 2020 research and innovation program under grant agreement No. 739593.

## A Detailed Simulation Results

In this section, we present detailed tables summarizing the outcomes of different intervention strategies under both *d*-step and stochastic mutation scenarios. The tables are quite straightforward, in each case we see the empirical probability 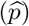 of the evolution with the corresponding standard deviation 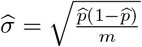.

### A.1 *d*-step Mutation

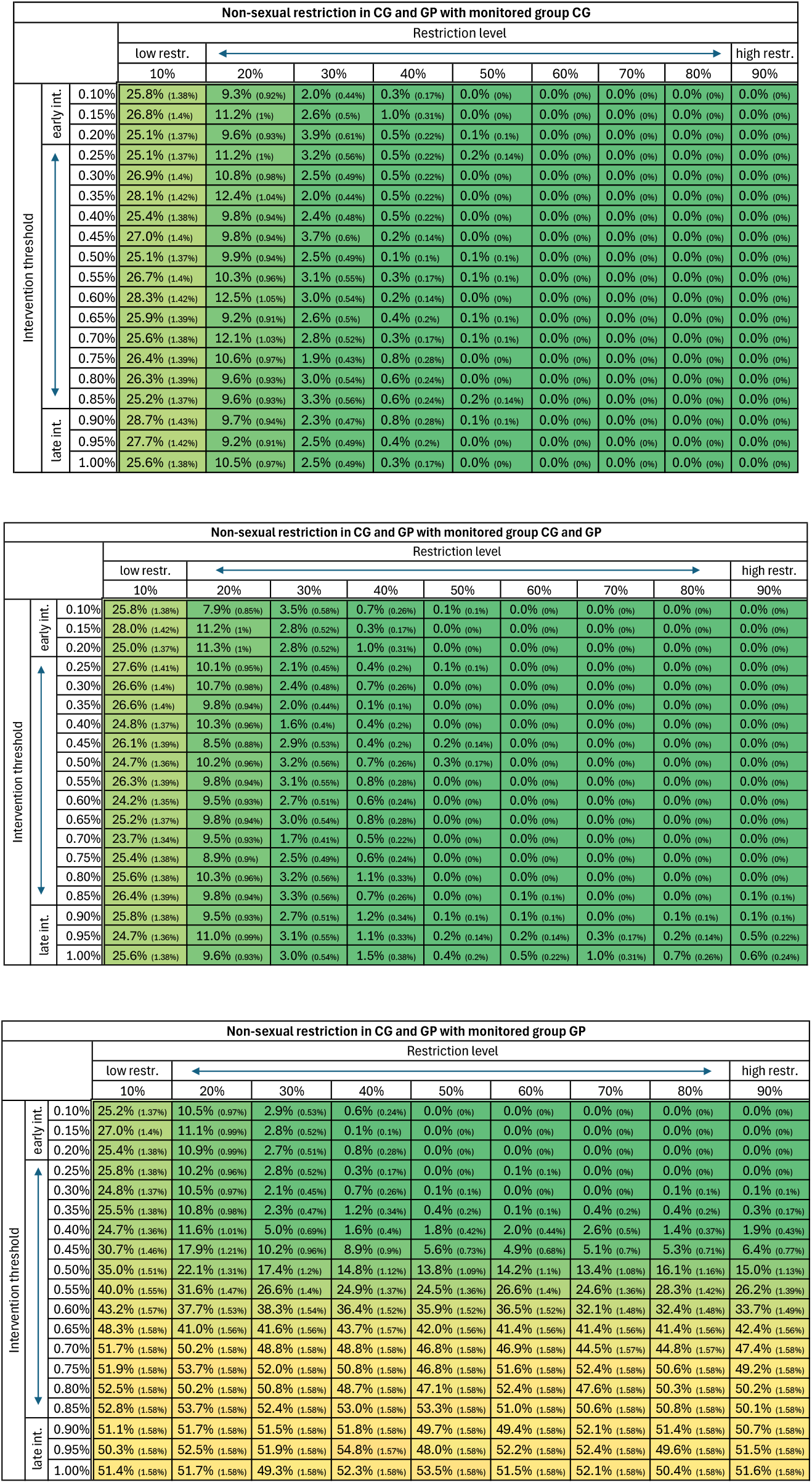
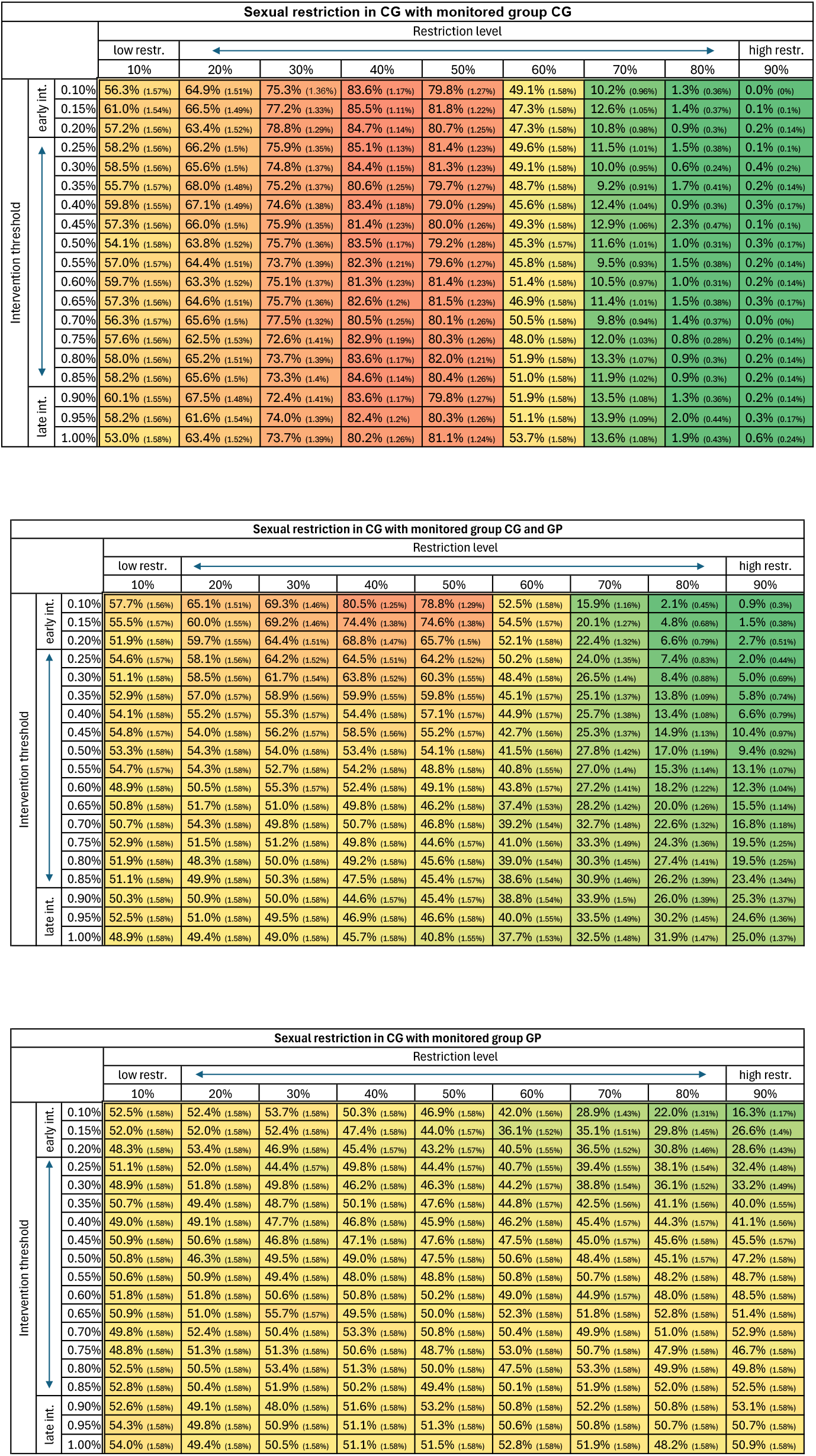

### A.2 Stochastic Mutation

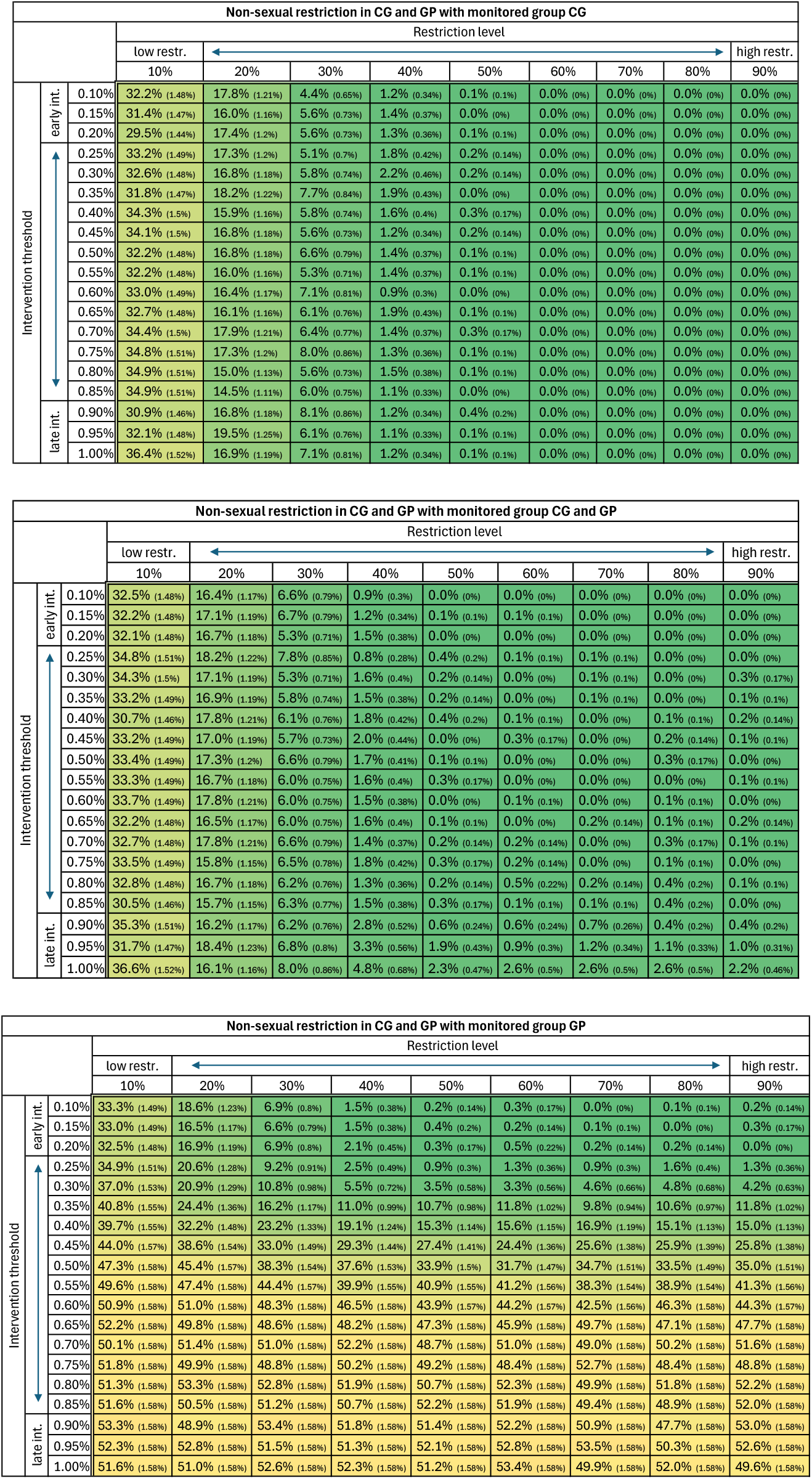
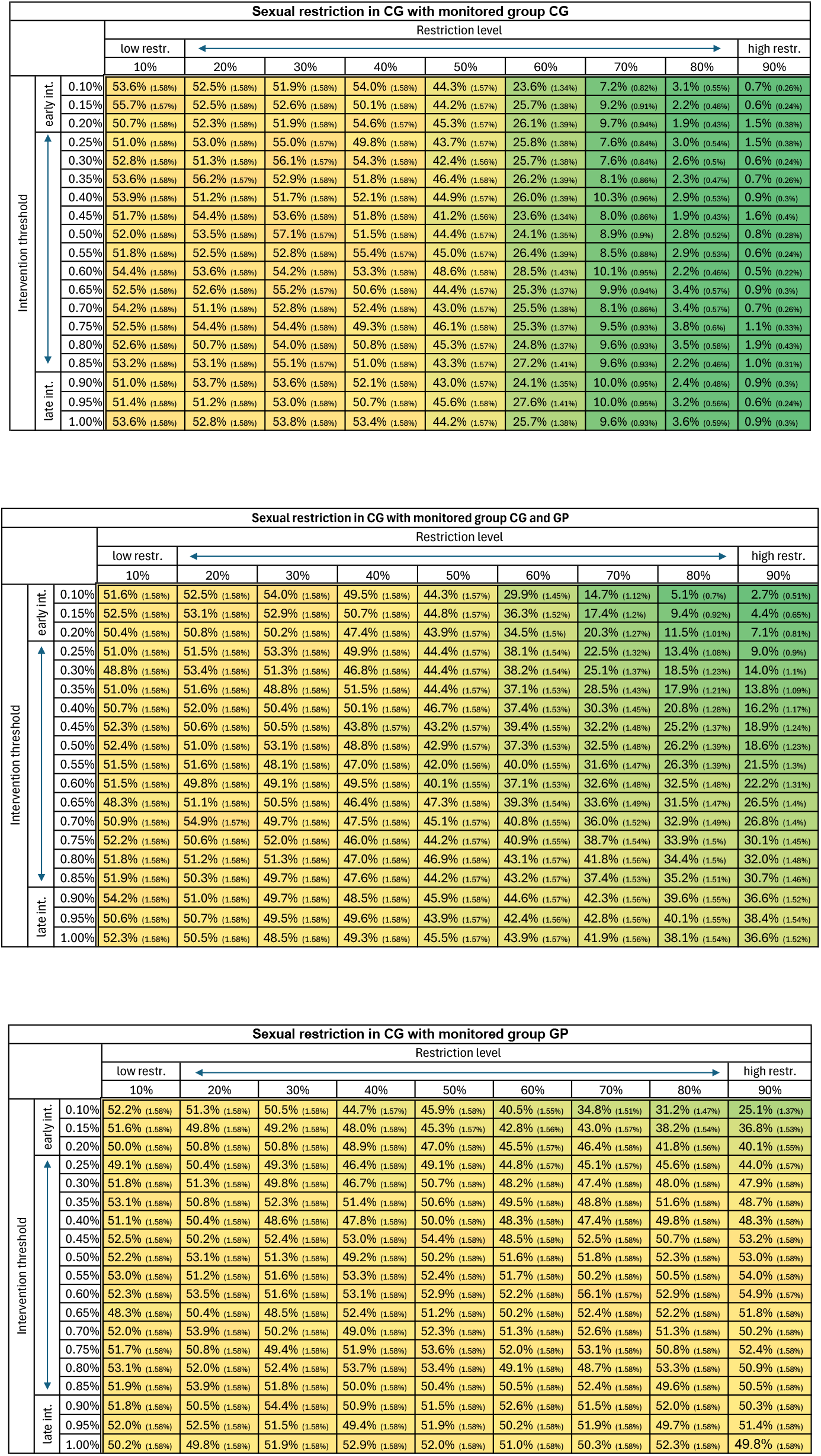

## B Transmission chain length

### B.1 *d*-step mutation

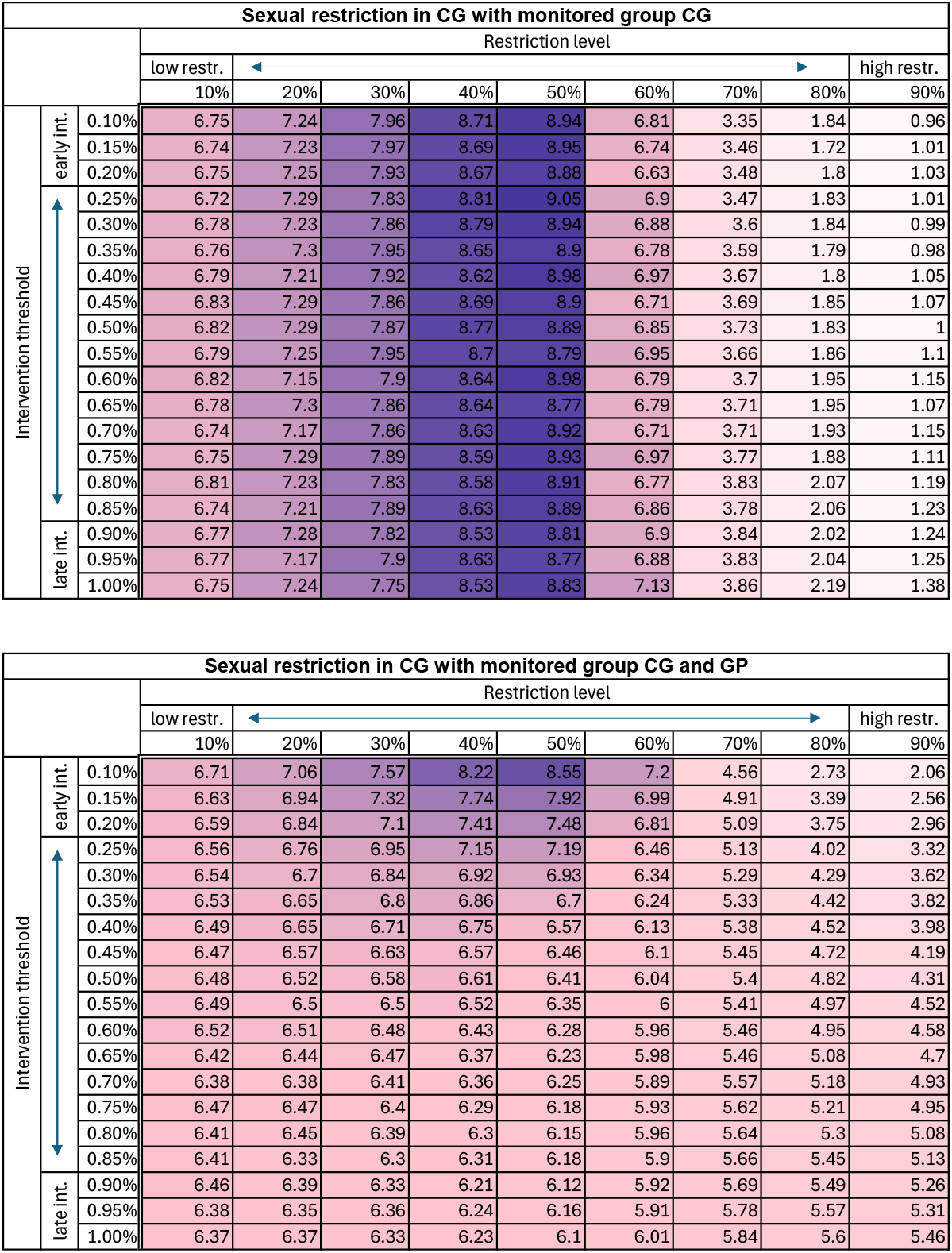

### B.2 Stochastic mutation

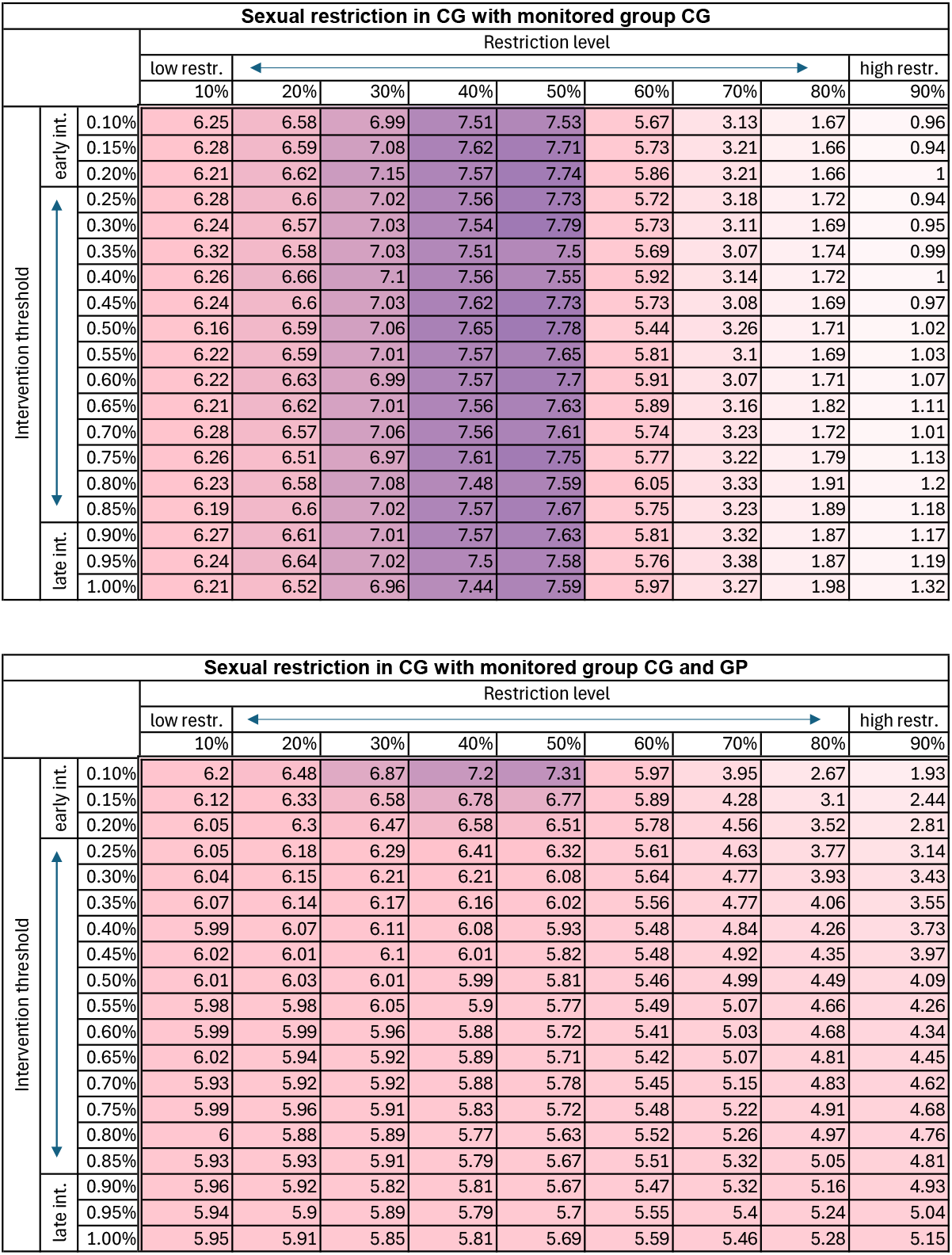

## C Pseudocode for Simulation Step

~~~
FUNCTION step()
      # Increment time by the time to the next action
      INCREMENT time by drawing from an exponential distribution
      with parameter equal to the sum of all action rates

      # Determine which group initiates the action
      SET action_group = randomly choose a group based on their
      aggregated action parameters
      # Determine the type of action within the chosen group
      SET action_type = randomly choose action type (exposure, infection,
      or recovery) based on the chosen group’s action parameters
      # Determine the specific individual within the chosen action type
      SET action_number = randomly choose an individual from the chosen
      group’s list of individuals for the specified action type
      # Execute the action
      IF action_type is exposure:
            # An exposed individual becomes infectious
            REMOVE individual from exposed list
            ADD individual to infectious list
     ELSE IF action_type is infection of GP:
            # An infectious individual from the chosen group infects a susceptible
            # individual in the general population
            SET infector = chosen infectious individual
            CALL mutate the infector’s non-sexual transmission probability
            DECREMENT susceptible count in general population
            CREATE new exposed individual in general population
            ADD new exposed individual to general population’s exposed list
    ELSE IF action_type is infection of CG:
          # An infectious individual from the chosen group infects a susceptible
          # individual in the core group
          SET infector = chosen infectious individual
          CALL mutate the infector’s non-sexual transmission probability
          DECREMENT susceptible count in core group
          CREATE new exposed individual in core group
          ADD new exposed individual to core group’s exposed list
    ELSE IF action_type is recovery:
          # An infectious individual recovers
          REMOVE individual from infectious list
          INCREMENT recovered count
    # Update tracking lists for plotting and analysis
APPEND current time to time list
FOR EACH group:
        APPEND current SEIR states to respective lists
        APPEND average R0 to R0 list
        APPEND maximum R0 to R0max list
        APPEND quartile R0 to R0quartile list
END FUNCTION
~~~

## D GitHub code

The codes performing our simulations and producing all the related outputs have been uploaded to GitHub. https://github.com/HCEMMScientificComputingACF/Mpox

